# A model-informed target product profile for population modification gene drives for malaria control

**DOI:** 10.1101/2024.08.16.24312136

**Authors:** Agastya Mondal, Héctor M. Sánchez C., John M. Marshall

## Abstract

As reductions in malaria transmission in sub-Saharan Africa stagnate, gene drive-modified mosquitoes represent one of the most promising novel tools for continued disease control. In order to advance from the laboratory to the field, gene drives will be assessed against target product profiles, planning tools that list minimum criteria products should satisfy as they progress through the development pipeline. Here, we use an eco-epidemiological model to investigate parameter values for population modification gene drives that satisfy two previously-discussed target outcomes: a 50% reduction in clinical malaria incidence for a duration (window-of-protection) of at least three years, and a time-to-impact of less than one year. We consider two African settings, Burkina Faso and Kenya, where gene drive mosquitoes are currently being researched, and consider three transmission intensities at each. For the gene drive product, we explore rates of homing and resistance allele generation, fitness costs associated with gene drive and non-functional resistance alleles, and the efficacy of the effector gene(s) at reducing mosquito-to-human transmission. We find that when the window-of-protection criterion is satisfied, the time-to-impact criterion also is. Target outcomes are most influenced by the fitness cost associated with the gene drive allele and effector gene efficacy. Resistance allele parameters are also highly influential on target outcomes, and determine how long the gene drive allele persists in the population after most available wild-type alleles have been cleaved. Low rates of functional resistance allele generation are preferred, while costly non-functional resistance alleles will allow the drive allele to outcompete them. Homing rates already achieved for *Anopheles* gene drives do not need to be improved upon. A conundrum exists whereby the most important product parameters for predicting field efficacy are those that can only be reliably measured in the field, which presents a challenge for assessment of product readiness.

## 1. Introduction

Malaria continues to pose a major public health burden throughout much of the world, especially in sub-Saharan Africa, where over 90% of cases and deaths occur [1]. Despite the wide-scale distribution of insecticide-based interventions and antimalarial drugs beginning in 2000, malaria persists at an unacceptably high level [1,2], and it is clear that new tools are needed for continued reductions in disease incidence and mortality. Two of the most promising novel tools at present are malaria vaccines and gene drive-modified mosquitoes. Gene drive approaches bias inheritance in favor of an introduced allele intended to spread through the mosquito population, and fall into two main categories: i) “population suppression,” whereby the introduced allele induces a fitness load or sex bias, reducing mosquito numbers, and ii) “population modification,” whereby the introduced allele disrupts pathogen transmission, reducing mosquito vector competence [3]. Candidate constructs for both approaches have been developed in the lab, most notably: i) a CRISPR-based system that targets the *doublesex* gene in *Anopheles gambiae*, the main African malaria vector, causing sterility in female homozygotes and inducing collapse of cage populations [4], and ii) CRISPR-based systems carrying dual antimalarial effector genes in *An. gambiae* and *Anopheles coluzzii*, that spread rapidly through cage populations [5]. Discussions regarding field trials of these systems are currently underway [6].

In order for gene drive mosquitoes to advance from the lab to the field, their characteristics should be assessed against target product profiles (TPPs), planning tools that provide a list of preferred characteristics and/or minimum criteria products must satisfy as they progress through the development pipeline. TPPs have recently been developed for a range of new malaria and vector control tools, including attractive targeted sugar baits (ATSBs) [7], long-acting injectable drugs [8], and malaria vaccines [9]. For mosquitoes engineered with low-threshold gene drive systems, developing and satisfying TPP criteria is particularly important given the potential for transgenes to become established in local mosquito populations following a release, and the expected difficulty of remediating these transgenes in the event of undesired outcomes or a shift in public opinion [10]. A draft TPP has been proposed for population modification gene drive products [11], and a workshop hosted by the Foundation for the National Institutes of Health (FNIH) discussed TPPs for gene drive products at length [12]. These preliminary discussions set a 20-50% reduction in clinical malaria incidence as a target outcome for a gene drive release, a rate of spread that would produce this impact within the time frame of a field trial (less than a year), and a duration of impact of at least 2-3 years. These target outcomes are subject to change pending wider stakeholder input.

Mathematical models will necessarily inform TPP criteria for gene drive mosquitoes as key target outcomes are primarily epidemiological (e.g., reductions in clinical malaria incidence) and can only be observed following a release [13]. Models are therefore needed to infer target parameter values for gene drive mosquito products, such as rates of homing and resistance allele generation, based on target epidemiological outcomes in the context of a given release scheme and setting. Fortunately, there has been a growth over the last 15 years in the field of malaria modeling, with several detailed models being developed that concisely describe malaria transmission dynamics in the mosquito vector and human host [14–16]. Concurrently, and particularly since the advent of CRISPR-based gene editing, several modeling frameworks have been developed to describe the population dynamics of mosquito genetic biocontrol tools [17–19]. Most relevant to TPPs, Leung *et al.* [20] used the EMOD malaria model to infer parameter values for population modification gene drive systems expected to eliminate malaria in low-to-moderate transmission settings in the Sahel, west Africa. The Imperial College London (ICL) malaria model has also been used to model epidemiological outcomes of population suppression and modification gene drives [21,22].

Here, we investigate parameter values for population modification gene drives that satisfy the epidemiological target outcomes outlined by James *et al.* [12], namely, a 50% reduction in clinical malaria incidence for a duration of at least three years, and a time-to-impact of less than a year. These outcomes represent the most demanding criteria specified by that expert group to reflect the fact that models, as a simplified representation of the real world, may overestimate the success of a tool that has not yet been field tested. To seed the population with the gene drive system, we simulate eight consecutive weekly releases of gene drive-modified *An. gambiae*, and consider this alongside existing interventions that include long-lasting insecticide-treated nets (LLINs), indoor residual spraying with insecticides (IRS), and artemisinin combination therapy drugs (ACTs). We consider two African settings, Burkina Faso and Kenya, with distinct seasonality and intervention profiles where gene drive mosquitoes are being actively researched, and consider three transmission intensities at each (high, medium and low). To characterize the gene drive construct, we consider parameters describing homing and resistance allele generation rates, the efficacy of the antimalarial effector gene(s), and fitness costs of gene drive and resistance alleles. We sample plausible ranges for each parameter and use the MGDrivE 3 model of mosquito genetic biocontrol [22], which incorporates a version of the ICL malaria transmission model [15,23], to predict epidemiological outcomes. Through analyzing the main drivers of the target outcomes, we provide an assessment of gene drive parameter values expected to satisfy TPP criteria, and discuss implications for future development efforts and lab and field measurements.

## 2. Methods

The impact of a population modification gene drive mosquito on malaria transmission will depend on both its product parameters and release setting. With this in mind, we explored the performance of gene drive at reducing malaria transmission for a range of parameters (rates of homing and resistance allele generation, transmission-blocking efficacy, and fitness effects, **Fig 1A**) in six distinct settings (sites resembling Burkina Faso and Kenya in terms of seasonal rainfall profile and intervention coverage, with three transmission intensities for each, **Fig 1B**). By randomly sampling a set of gene drive parameters for each setting, a set of simulations was defined and run (**Fig 1C-D**) and outcomes of interest were extracted from the simulation output (**Fig 1E**). An emulator was then trained with data from the simulation bank to predict disease outcomes given input parameters [24]. Finally, the emulator was employed to conduct a sensitivity analysis and determine gene drive parameter ranges that satisfy TPP criteria (**Fig 1F**).

**Fig 1.**
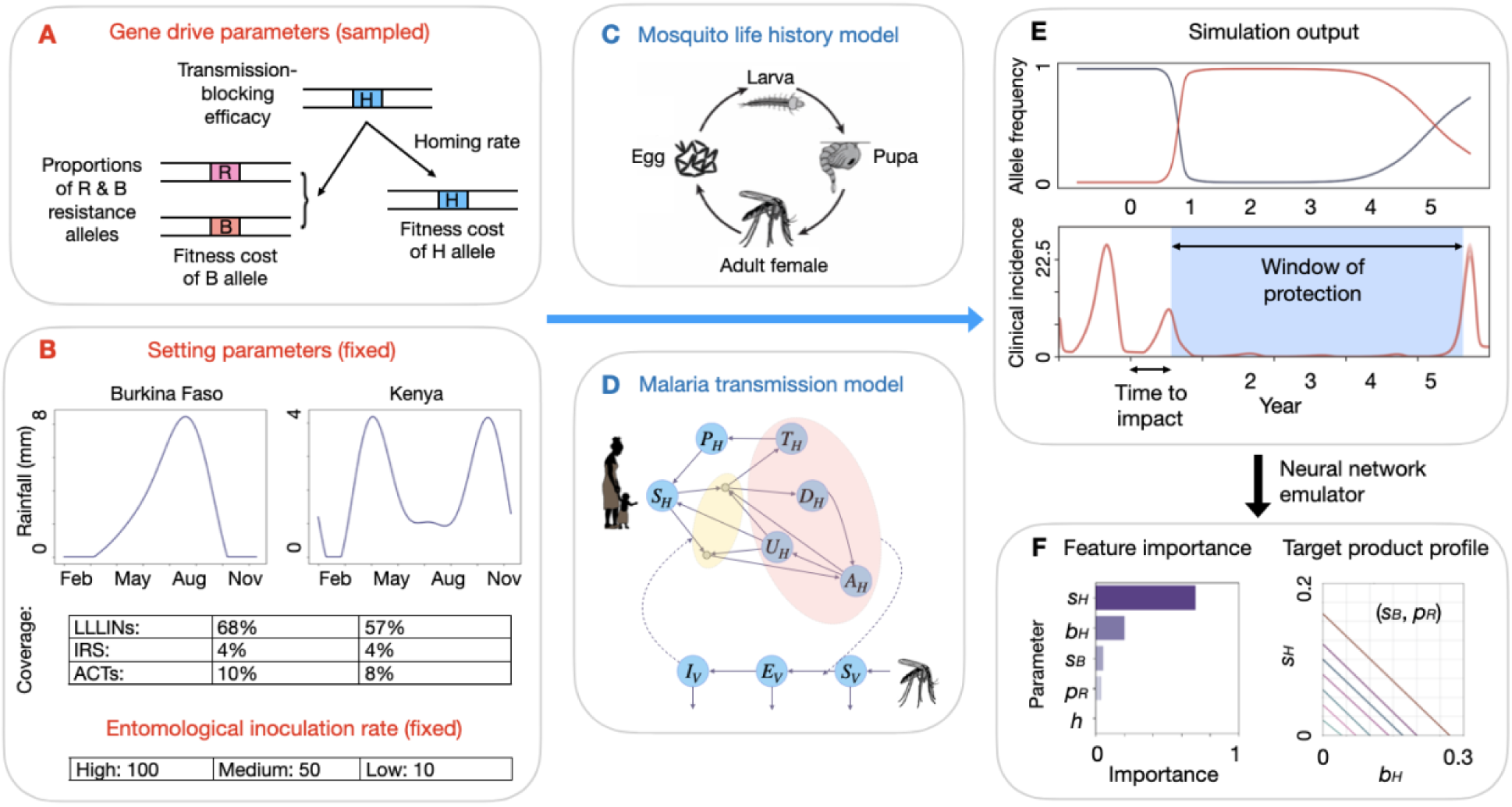
Workflow diagram to assess target product profile of gene drive-modified mosquitoes. **(A)** We consider a population modification gene drive system and antimalarial effector gene(s) with defined transmission-blocking efficacy (*b_H_*). When present in a heterozygote, the gene drive allele (H) cleaves a wild-type allele in the germ line, either converting it into an H allele through homology-directed repair (at a homing rate, *h*), or into a resistance allele that is either in-frame/cost-free (R, with probability *p_R_*), or out-of-frame/otherwise costly (B). Fitness costs are defined for H and B alleles (*s_H_* and *s_B_*, respectively). For each gene drive parameter, a distribution of values are defined and sampled. **(B)** Simulations are performed for two settings (Burkina Faso and Kenya) and three transmission settings (entomological inoculation rates of 100, 50 and 10 infective bites per person per year). Settings are defined by their seasonal rainfall profile and coverage of currently-available tools including long-lasting insecticide-treated nets (LLINs), indoor residual spraying with insecticides (IRS), and artemisinin combination therapy drugs (ACTs). **(C)** Simulations are run using the MGDrivE 3 modeling framework [22], which includes modules for gene drive inheritance, mosquito life history and malaria epidemiology. Mosquito life history is modeled according to an egg-larva-pupa-adult life cycle, with environmental carrying capacity for larvae modulated by recent rainfall. **(D)** Malaria transmission is modeled according to the Imperial College London malaria model [15,25]. Here, humans progress from susceptible (*S_H_*) to either symptomatic or asymptomatic infection. Humans who develop a symptomatic infection and are either treated (*T_H_*) or diseased and untreated (*D_H_*). Treated humans advance to a prophylactic protection state (*P_H_*) and eventually become susceptible again. Untreated symptomatic humans develop successively lower-density infections, from symptomatic to asymptomatic but detectable by rapid diagnostic test (RDT) (*A_H_*) to asymptomatic and undetectable by RDT (*U_H_*). In parallel, adult female mosquitoes progress from susceptible (*S_V_*) to exposed/latently infected (*E_V_*) to infectious for malaria (*I_V_*). **(E)** For each sampled parameter set and setting, gene drive allele frequencies and clinical malaria incidence are recorded for six years. Window-of-protection (WOP, the duration for which clinical incidence is below 50% its seasonal mean) and time-to-impact (TTI, the time from initial release to clinical malaria incidence falling to 50% its seasonal mean) are recorded as outcomes of interest. **(F)** Neural network emulators are trained using gene drive parameter values (*b_H_*, *h*, *p_R_*, *s_H_* and *s_B_*) and outcomes (WOP and TTI) for each setting. Emulators are then used to calculate importance of each parameter in predicting WOP and TTI, and to infer regions of gene drive parameter space that satisfy WOP >3 years and TTI <1 year.

### Gene drive and malaria transmission model

We used the MGDrivE 3 framework [22] to simulate releases of *An. gambiae* mosquitoes engineered with a population modification gene drive system and linked antimalarial effector gene. MGDrivE 3 is a stochastic, population-based model that simulates the population dynamics of mosquito genetic control tools and their entomological and epidemiological implications. The framework includes: i) an inheritance module that describes the distribution of offspring genotypes for given maternal and paternal genotypes, ii) a life history module that describes the development of mosquitoes from egg to larva to pupa to adult (**Fig 1C**), and iii) an epidemiology module that describes reciprocal pathogen transmission between mosquitoes and humans (**Fig 1D**). A landscape module that describes the distribution and movement of mosquitoes through a metapopulation is also included, but was not utilized for this analysis. Seasonality in mosquito density is incorporated through time-dependent mosquito bionomic parameters, which are responsive to environmental data. In the present analysis, data on recent rainfall (**Fig 1B**) modulate the carrying capacity of the environment for larvae, which in turn impacts adult mosquito density and malaria transmission.

The MGDrivE 3 framework is linked to an adapted version of the Imperial College London (ICL) malaria transmission model [15,25] (**Fig 1D**). The ICL malaria model has been fitted to extensive data sets throughout sub-Saharan Africa and captures important details of malaria transmission, including symptomatic and asymptomatic infection, variable parasite density and superinfection in humans, human age structure, mosquito biting heterogeneity, several forms of immunity, and antimalarial drug therapy and prophylaxis. Treatment coverage with ACTs is captured within the ICL malaria model, while coverage with vector control interventions, such as LLINs and IRS, is captured within the mosquito life history module of MGDrivE 3. Here, mosquito life history parameters are modified to reflect the fact that LLINs and IRS increase the mortality rate and decrease the biting rate of adult mosquitoes, and also decrease the egg-laying rate by virtue of extending the gonotrophic cycle [15,26]. Gene drive interventions are therefore modeled in the context of existing coverage with ACTs, LLINs and IRS (**Fig 1B**). The ICL malaria model permits monitoring a variety of health outcomes, e.g., clinical disease incidence, prevalence and mortality by age group. We focused on all-ages clinical incidence of malaria for this analysis.

### Gene drive product parameters and release scheme

MGDrivE 3 allows for flexible specification of genetic constructs and release schemes. To model a generic population modification gene drive, we consider an inheritance cube including a homing allele (H), wild-type allele (W), and two varieties of homing-resistant alleles, one that is in-frame and cost free, sometimes referred to as “functional” (R), and another that is out-of-frame or otherwise costly (e.g., a large in-frame deletion), sometimes referred to as “non-functional” (B) (**Fig 1A**). Inheritance cubes were introduced in the first version of MGDrivE [18] and describe the distribution of offspring genotypes given parental genotypes. This inheritance cube has been used to model population modification gene drives in *Anopheles stephensi* [27,28], *An. gambiae* [22,29], and other species [30,31]. In this cube, Mendelian inheritance rules apply at the gene drive locus, with the exception that, for adults heterozygous for the H and W alleles, a proportion, *c*, of W alleles are cleaved, while a proportion, 1−*c*, remain as W alleles. Of those that are cleaved, a proportion, *h*, are subject to accurate homology-directed repair (HDR) and become H alleles, while a proportion, 1−*h*, become resistance alleles. Of those that become resistance alleles, a proportion, *p_R_*, become R alleles, while the remainder, 1−*p_R_*, become B alleles. Each of these parameters may differ depending on the sex of the HW individual, but we did not consider sex-specific parameters in this analysis to reduce dimensionality. Fitness effects may be associated with any genotype, but here we considered just two fitness parameters to reduce dimensionality: *s_H_*, which represents reductions in female fecundity and male mating competitiveness associated with being homozygous for the H allele, and *s_B_*, which represents the same costs associated with being homozygous for the B allele. Fitness costs are assumed to be additive. Finally, the H allele is assumed to be linked to an antimalarial effector gene, and *b_H_* represents the probability of mosquito-to-human infection for a mosquito having at least one copy of the H allele.

For each gene drive parameter, a distribution of values is defined and sampled from to inform the TPP analysis (**Table 1**). Given perfect or near-perfect cleavage of W alleles in HW heterozygotes for recent *An. gambiae* gene drive constructs [5,32], we assume a cleavage rate, *c*, of 1. Probabilities of accurate HDR, *h*, tend to be high for *Anopheles* gene drives, e.g., ∼0.98 for AcTP13 in *An. coluzzii* females and males [5], and 0.96 and 0.98 for AgNosCd-1 in *An. gambiae* females and males, respectively [32]. We explore a wider range of homing rates between 0.8 and 1.0 for this TPP analysis. The proportion of resistance alleles that are in-frame/cost-free (R), *p_R_*, varies depending on the construct, and whether the resistance allele is formed in the parental gamete (e.g., via non-homologous end-joining) or embryo (following maternal deposition of Cas) [33]. For AsMCRkh2 in *An. stephensi*, models fitted to data were consistent with 0.005 of generated resistance alleles being R in gametes and 0.22 being R in embryos [27]. For Rec*kh* in *An. stephensi*, fitted models suggested the proportion of R resistance alleles to be 0.17 in both gametes and embryos [28]. These estimates are consistent with about a third of mutations preserving the reading frame, and some fraction of those being cost-free (i.e., R). We define an exponential distribution for *p_R_* with a mean of 0.11 to reflect 86% of values being between 0 and 0.22. To reduce dimensionality, we ignore maternal deposition of Cas in this analysis, as model analyses of the AgTP13 and AcTP13 constructs support the conclusion that gene drive outcomes are insensitive to the stage of resistance allele generation [5].

**Table 1.** Gene drive and setting parameters used for implementation simulations.

|  | Parameter | Value(s) |  | Reference(s) |
| --- | --- | --- | --- | --- |
| **Gene drive mosquito product** | Species | *An. gambiae* |  |  |
| | DNA cleavage rate ($c$) | 1 | | [5,32] |
| | Probability of accurate homology-directed repair given cleavage ($h$) | ~Unif [0.8-1] | | [5,27,28,32] |
| | Proportion of resistant alleles that are in-frame/cost-free ($p_R$) | ~Exp (mean = 0.11) | | [27,28] |
| | Reduction in female fecundity & male mating competitiveness in HH individuals ($s_H$) | ~Unif [0-0.4] | | |
| | Reduction in female fecundity & male mating competitiveness in BB individuals ($s_B$) | ~Unif [0-0.4] | | |
| | Heterozygosity of fitness costs ($h_s$) | 0.5 | | |
| | Probability of mosquito-to-human infection ($b_H$) | ~Exp (mean = 0.16) | | [5] |
| | Probability of maternal deposition of Cas ($m_D$) | 0 | | [5] |
|  |  | Burkina Faso | Kenya |  |
| **Setting** | Rainfall seasonality | Unimodal (Fig 1) | Bimodal (Fig 1) | [34] |
| | Long-lasting insecticide-treated net coverage ($\theta_{ITN}$) | 68% | 57% | [35] |
| | Indoor residual spraying coverage ($\theta_{IRS}$) | 4% | 5% | [35] |
| | Artemisinin-based combination therapy coverage ($f_T$) | 10% | 8% | [35] |
| | Entomological inoculation rate ($\varepsilon$) | (10, 50, 100) per person per year | | [36] |

Values represent probabilities/proportions, percentages, or values with other defined units. Unif [ $a$ - $b$ ] represents a uniform distribution with minimum, $a$ , and maximum, $b$ . Exp (mean = $x$ ) represents an exponential distribution with mean, $x$ .

Fitness costs corresponding to gene drive alleles are somewhat hypothetical at present as they are yet to be estimated in the field, and laboratory estimates are of limited relevance. For example, laboratory studies of AcTP13 in *An. coluzzii* found that the H allele has a fitness advantage over the W allele [5], which is unlikely to be realized in the field. In other laboratory experiments, studies reveal approaches to minimize fitness costs. For example, for AsMCRkh2 in *An. stephensi*, fitted models were consistent with a fitness cost on H of 0.08 per allele, and a fitness cost on B of 0.18 per allele [27], but when the target *kynurenine hydroxylase* gene was recoded in the Rec*kh* construct, fitted models suggested that a fitness cost on H was no longer present [28]. Given the uncertainty surrounding fitness costs in the field, we consider additive fitness costs between 0 and 0.2 per allele for both H and B alleles; or equivalently, homozygous fitness costs, *s_H_* and *s_B_*, between 0 and 0.4 for HH and BB individuals, respectively. The transmission-blocking efficacy of potential antimalarial effector genes is also quite hypothetical at present, as measurements have been made of parasite life stages in the vector; but not of onward transmission to human hosts [5,37]. The probability of mosquito-to-human infection for a wild-type mosquito, *b*, is estimated at 0.55 [38]. Antimalarial effectors in the AgTP13 and AcTP13 constructs reduce sporozoite density in mosquito salivary glands and, under various assumptions about sporozoite densities required for infection, produce mosquito-to-human infection probabilities, *b_H_*, between 0 and 0.32 [5]. We define an exponential distribution for *b_H_* with a mean of 0.16 to reflect 86% of values being between 0 and 0.32.

Finally, we consider a release scheme consisting of eight consecutive weekly releases of 20,000 HH male *An. gambiae* released at the beginning of the rainy season in each setting. The wild *An. gambiae* population size is dynamic, and is calibrated to produce the specified EIR in each country setting, so the initial release frequency varies depending upon the country setting, transmission intensity and timing of release. In Burkina Faso, with releases timed for the beginning of seasonal rains, these releases represent an initial population frequency of 0.16 (low EIR), 0.011 (medium EIR) or 0.0024 (high EIR), and in Kenya of 0.28 (low EIR), 0.017 (medium EIR) or 0.0045 (high EIR).

### Simulated settings, seasonality and other interventions

Using MGDrivE 3, which incorporates the ICL malaria model, we simulate two African settings, Burkina Faso, where gene drive mosquitoes are being actively researched, and consider three transmission settings for each. Rainfall is a major driver of *An. gambiae* population dynamics, and these two countries have distinct seasonal rainfall patterns, Burkina Faso has a single rainy season from May through September, while Kenya has a “long rains” season from March through May and a “short rains” season from October through December. **Fig 1B** depicts smoothed rainfall profiles for each country derived using the “umbrella” R package [39]. This package fits a mixture of sinusoids to daily rainfall data from the CHIRPS (Climate Hazards Group Infrared Precipitation with Station) database [34], here representing the three years between January 1st, 2017 and December 31st, 2019. In MGDrivE 3, recent rainfall modulates the carrying capacity of the environment for larvae via a mathematical relationship from White *et al.* [40]. To maintain a persistent *An. gambiae* population throughout the year, we assume larval carrying capacity in the dry season is 5% that of the peak rainy season, qualitatively consistent with entomological data from Burkina Faso [41] and Kenya [42] concluding that vector breeding sites are substantially less abundant during the dry season.

Simulated settings are also characterized by their coverage of existing interventions that include LLINs, IRS and ACTs, which are modeled alongside gene drive releases. Country-specific coverage levels for these interventions were obtained from the Malaria Atlas Project [35] and are included in **Table 1**. Coverage levels with LLINs and IRS modify the mortality, biting and egg-laying rates of adult mosquitoes based on the model of Le Menach *et al.* [26]. Finally, we consider three transmission intensities for each setting, EIRs (entomological inoculation rates) of 100 (high), 50 (medium), and 10 (low) infectious bites per person per year. Time-varying mosquito density was scaled to produce each EIR prior to the gene drive release, thus accounting for seasonal rainfall profiles and coverage with existing interventions. Simulations were run for a human population size of 1,000, consistent with a medium-sized Burkinabe or Kenyan village. Additional parameter values describing mosquito bionomics, vector control and malaria epidemiology not listed in **Table 1** are available in **S1 Table**.

### Target outcomes and metrics

The primary metric for this TPP analysis is model-predicted all-ages clinical incidence of malaria, i.e., the number of new symptomatic malaria cases per day across all age groups. For each simulation, this is generated as a time-series, and two target outcomes are derived: i) window-of-protection (WOP), which measures the duration for which clinical incidence is below 50% its seasonal mean, and ii) time-to-impact (TTI), which measures the time from initial release to clinical malaria incidence falling to 50% its seasonal mean (**Fig 1E**). Calculation of clinical incidence at time *t*, *Inc*(*t*), follows from the ICL malaria model [15,25] (**Fig 1D**),

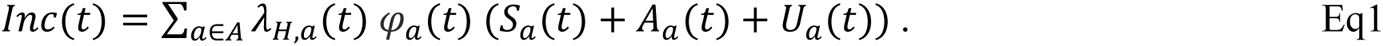

Here, *λ_H_*_,*a*_(*t*) represents the force of infection on humans (probability of infection per person per unit time) for age group *a* at time *t*, *φ_a_*(*t*) represents the probability of acquiring clinical disease upon infection for age group *a* at time *t* (this depends on age-specific immunity levels in the population), and *S_a_*(*t*), *A_a_*(*t*) and *U_a_*(*t*) represent the number of people in age group *a* who are either susceptible, asymptomatic but detectable by rapid diagnostic test (RDT), or asymptomatic and undetectable by RDT, respectively, at time *t*.

As secondary metrics for the TPP analysis, we also calculated all-ages malaria prevalence and malaria-induced mortality over time, calculating WOP and TTI outcomes for each. Malaria prevalence, also referred to as the *Plasmodium falciparum* parasite rate (*PfPR*), refers to the proportion of the human population that harbors the malaria pathogen, regardless of symptoms or treatment status. For the ICL malaria model [15,25], this is given by,

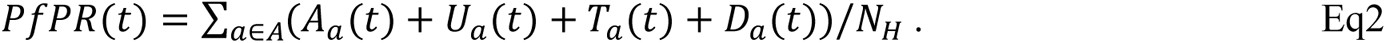

Here, *N_H_* represents the total human population size, *A_a_*(*t*) and *U_a_*(*t*) are as previously defined, and *T_a_*(*t*) and *D_a_*(*t*) represent the number of people in age group *a* who are symptomatically infected and either treated or untreated (diseased), respectively, at time *t*. Finally, for the ICL model [15,25], malaria-induced mortality is proportional to the incidence of severe malaria at time *t*, and is given by,

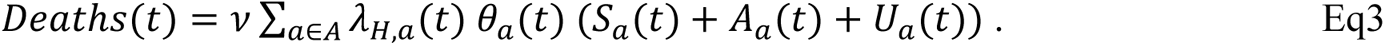

Here, *θ_a_*(*t*) represents the probability of acquiring severe disease upon infection for age group *a* at time *t* (this depends on age-specific immunity levels in the population), and *ν* represents the probability of death for a case of severe disease. The derivation and parameterization of this formula is provided in Griffin *et al.* [25].

### Neural network emulator to predict epidemiological impact

In order to rapidly search gene drive parameter space to infer the impact of product parameters on epidemiological target outcomes, we used a database of MGDrivE 3 simulations to train a neural network emulator for each EIR and country setting. To generate the simulation database, we sampled 3,000 gene drive parameter sets per setting/EIR using a Latin hypercube sampling scheme and the parameter distributions specified in **Table 1**. This provided a total of 18,000 simulations (3,000 parameter sets x 2 settings x 3 EIRs). Each simulation was run ten times, with the mean outcome being recorded, to avoid outliers given the stochastic nature of the model. Input parameters to the neural network for each setting/EIR are those listed in **Table 1**, i.e.: i) the homing rate, ii) proportion of resistance alleles that are in-frame/cost-free, iii) fitness cost on HH individuals, iv) fitness cost on BB individuals, and v) probability of mosquito-to-human transmission for mosquitoes having the effector gene. Output parameters are the WOP and TTI for the three outcome metrics described in the previous section -clinical incidence of malaria, malaria prevalence, and malaria-induced mortality. The emulators were trained on 90% of the simulation data and evaluated on a 10% hold-out test set. Python packages “tensorflow” [43] and “keras” [44] were used to build each emulator. The emulation utilized a simple feed-forward architecture with {32, 32} nodes in each fully-connected, hidden layer, and rectified linear unit activation functions between each hidden layer.

### Feature importance and target product profile

To assess the relative importance of each gene drive product parameter on the WOP and TTI target outcomes, we calculated their “feature importance” using the permutation feature importance method (**Fig 1F**). Permutation feature importance measures the decrease in a model score when a single feature value is shuffled, decoupling it from its outcome. The decrease in model score conveys the relative importance of the shuffled feature on the model’s outcome on a 0-1 scale. We used the Python package “scikit-learn” [45] to calculate feature importance scores. Having identified the most important gene drive product parameters, we next used the emulator to identify regions of parameter space that satisfy TPP criteria (**Fig 1F**), i.e., a WOP greater than three years, and a TTI less than one year. TPP criteria were identified for each setting/EIR combination, and for each of the three outcome metrics -clinical malaria incidence, malaria prevalence and malaria-induced mortality.

## 3. Results

### Target outcomes are most influenced by gene drive allele fitness cost and effector gene efficacy

We found a consistent ordering of gene drive product parameters when assessing “feature importance” for both WOP and TTI outcomes for the clinical malaria incidence metric, regardless of country setting, EIR or target outcome (**Fig 2**). The most influential parameter in all cases was the fitness cost associated with the intact gene drive allele, *s_H_*. Following this was the mosquito-to-human infection probability, *b_H_*, for mosquitoes having the gene drive allele and effector gene(s). Notably, the homing rate, *h*, or rate of accurate HDR given cleavage, was consistently the least influential of all product parameters explored. This is likely due to the high inheritance bias associated with the gene drive allele for the full range of homing rates explored, i.e., from 0.8 to 1, despite the lower bound of this range being substantially lower than values published for recently engineered population modification gene drives in *Anopheles* [4,5,27,28,32]. Moderate influence was imposed by resistance allele parameters: the fitness cost associated with out-of-frame or otherwise costly resistance B alleles, *s_B_*, followed by the proportion of generated resistance alleles that are in-frame and cost-free, *p_R_*.

**Fig 2.**
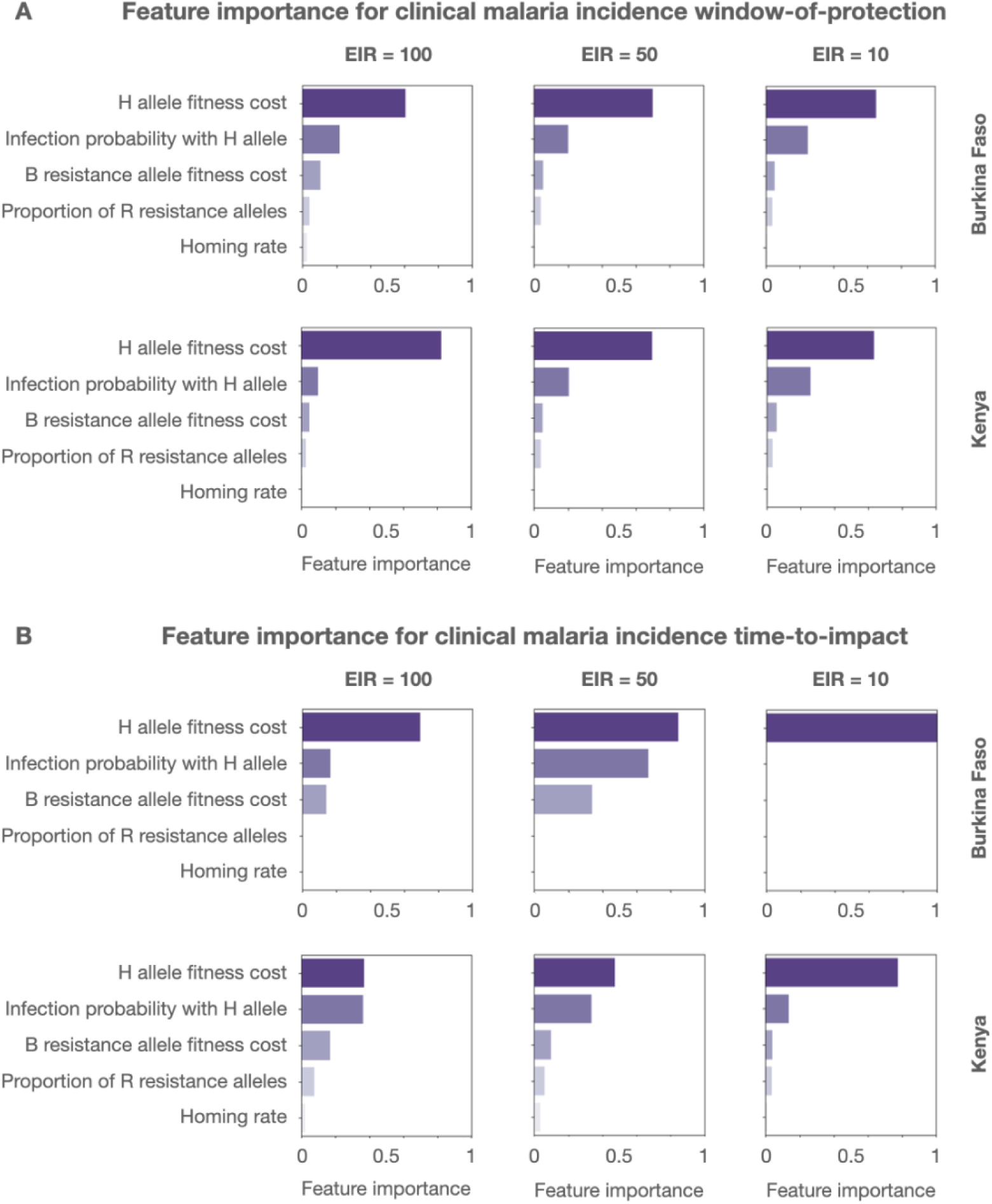
Feature importance of gene drive product parameters. Permutation feature importance values are depicted for gene drive product parameters for two country settings (Burkina Faso and Kenya), three transmission intensities (entomological inoculation rates (EIRs) of 100, 50 and 10 per person per year), and two target outcomes - **(A)** window-of-protection (i.e., the duration for which clinical incidence is below 50% its seasonal mean), and **(B)** time-to-impact (i.e., the time from initial release to clinical malaria incidence falling to 50% its seasonal mean). Parameters explored include: i) the fitness cost associated with being homozygous for the gene drive (H) allele, ii) the probability of mosquito-to-human transmission for mosquitoes having the H allele with linked antimalarial effector gene(s), iii) the fitness cost associated with being homozygous for the out-of-frame or otherwise costly B resistance allele, iv) the proportion of generated resistance alleles that are in-frame and cost-free (R), and v) the homing rate, or rate of accurate homology-directed repair given cleavage. Permutation feature importance is calculated on a 0-1 scale.

While the ordering of gene drive product parameters with respect to feature importance is consistent, their relative magnitudes vary depending on country setting, EIR and target outcome (**Fig 2**). H allele fitness cost, for instance, has more than double the feature importance magnitude than the next most influential parameter when considering the WOP outcome for clinical malaria incidence, regardless of country setting or EIR; however for the TTI outcome, mosquito-to-human infection probability is a close second in feature importance for high EIRs in both countries and medium EIRs in Kenya. For the TTI outcome, there is also a consistent increase in magnitude of feature importance for resistance allele parameters (B allele fitness cost and proportion of R resistance alleles) with declining EIR; however for the WOP outcome, the feature importance of these parameters remains small across all country settings and EIRs. Similar patterns are seen for the WOP and TTI outcomes for malaria prevalence and malaria-induced mortality metrics (see **Figs S1-S2** in **S1 Text**).

### Trade-offs between gene drive allele fitness cost and effector gene efficacy

To characterize regions of gene drive parameter space that satisfy TPP outcome criteria, we visualize the WOP for a >50% reduction in clinical malaria incidence as it varies with the four most influential product parameters, *s_H_*, *b_H_*, *s_B_* and *p_R_*, for both country settings and three EIRs for each (**Fig 3**). We excluded the homing rate parameter, *h*, from this analysis since all proposed outcome criteria were found to be least sensitive to its value within the range 0.8 to 1. We set the default value for *h* to 0.95, as this is a relatively high homing rate that is exceeded for all constructs, in both female and male lineages, that are currently being considered for malaria vector control [4,5,32].

**Fig 3.**
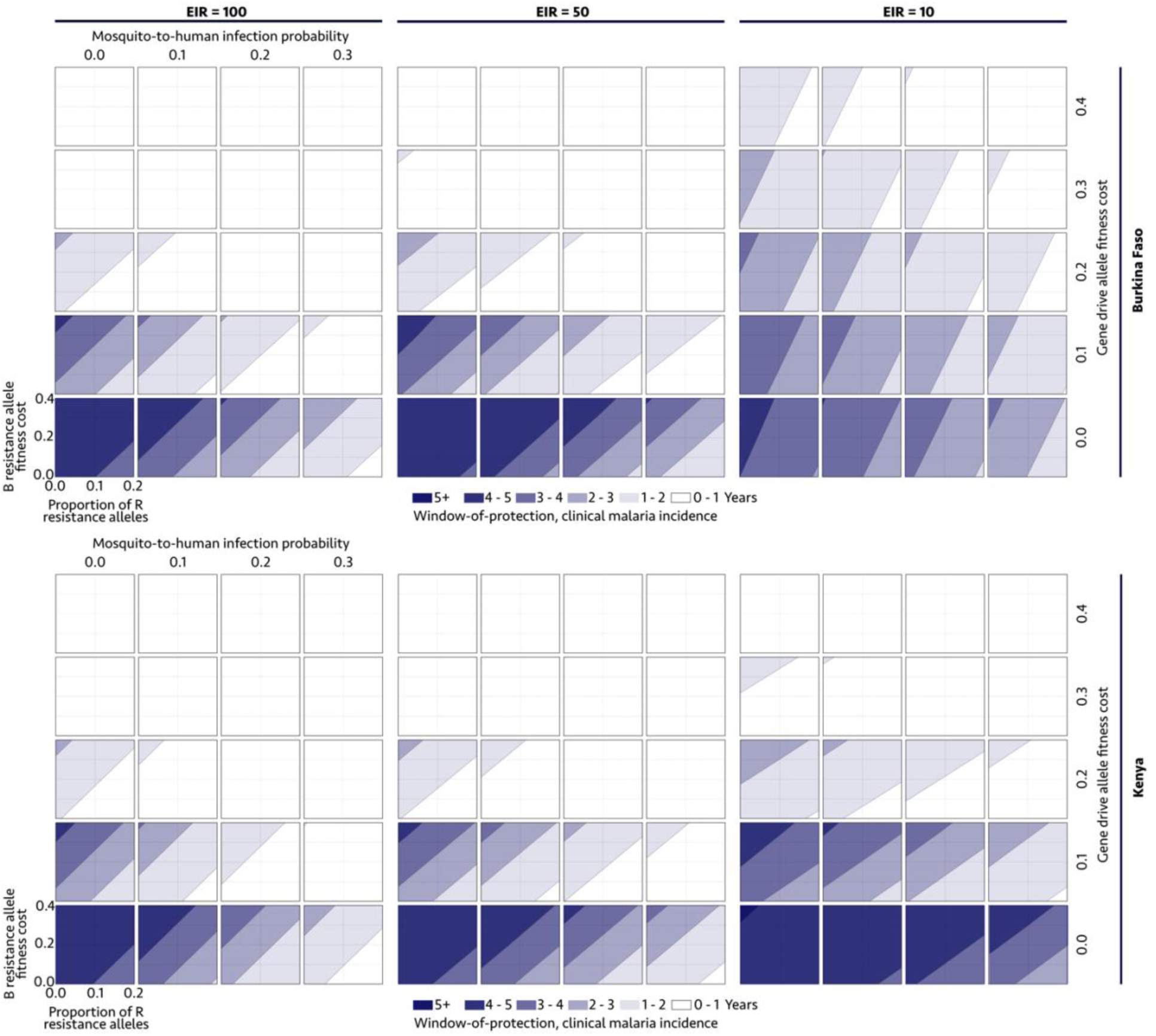
Gene drive parameter space satisfying a >50% reduction in clinical malaria incidence for defined durations. Windows-of-protection (WOPs) are depicted for two country settings (Burkina Faso and Kenya, defined by their seasonal profile in **Fig 1B** and intervention coverage profile in **Table 1**) and three transmission settings (entomological inoculation rates, or EIRs, of 100, 50 and 10 infective bites per person per year). Gene drive parameters explored include: i) the fitness cost associated with being homozygous for the gene drive (H) allele, ii) the probability of mosquito-to-human transmission for mosquitoes having the H allele, iii) the fitness cost associated with being homozygous for the out-of-frame or otherwise costly B resistance allele, and iv) the proportion of generated resistance alleles that are in-frame and cost-free (R). The homing rate parameter is fixed at 0.95, since proposed outcome criteria were found to be least sensitive to its value within a feasible range.

Results in **Fig 3** suggest a trade-off between gene drive allele fitness cost, *s_H_*, and effector gene efficacy, *b_H_*, for satisfying clinical malaria incidence WOP criteria. For most settings (country and EIR), there are scenarios in which the WOP exceeds three years for an infection probability of *b_H_*≤0.3 in the absence of a gene drive fitness cost, and for an infection probability *b_H_*≤0.1 in the presence of a fitness cost of *s_H_*≤0.1. If the target outcome is relaxed to a WOP exceeding two years, for most settings (country and EIR), there are scenarios in which this target outcome is satisfied for an infection probability of *b_H_*≤0.3 in the absence of a gene drive fitness cost, for an infection probability *b_H_*≤0.2 in the presence of a fitness cost of *s_H_*≤0.1, and for perfect transmission-blocking in the presence of a fitness cost of *s_H_*≤0.2. That is, higher gene drive fitness costs can be tolerated for higher effector gene efficacies. Results for moderate-to-high EIRs (50-100 infective bites per person per year) are relatively consistent; however, slightly higher infection probabilities can be tolerated for the same gene drive allele fitness cost for low-to-moderate EIRs (10-50 infective bites per person per year).

Gene drive parameter values that satisfy WOP criteria for malaria-induced mortality reflect those for clinical malaria incidence (**Fig S3** in **S1 Text**). This is partly due to the fact that malaria-induced mortality is calculated as a proportion of the incidence of severe malaria (Eq 3), which differs from the incidence of clinical malaria (Eq 1) by only one term. That said; WOP criteria for malaria-induced mortality are satisfied for a slightly wider range of gene drive allele fitness and infection probability parameters, possibly due to the fact that reductions in severe disease occur sooner than reductions in clinical disease following an intervention-induced reduction in transmission. WOP criteria based on the malaria prevalence metric (Eq 2) are more restrictive on admissible gene drive parameter space (**Fig S4** of **S1 Text**), likely due to the fact that untreated malaria infections can last for months to years [46], and consequently, the greatest reduction in malaria prevalence may not be seen until intervention impact begins to wane due to resistance allele spread in many cases.

Results in **Fig 4** confirm that the TTI outcome criterion (a TTI of less than one year) is satisfied for all regions of gene drive parameter space that satisfy the WOP outcome criterion for the clinical malaria incidence metric. This is helpful to realize as it means that, in developing a model-informed TPP, we can focus entirely on regions of gene drive parameter space that satisfy the WOP outcome criterion. Nevertheless, **Fig 4** provides additional information on how the TTI varies with gene drive and setting parameters. Limiting consideration to gene drive parameters that satisfy the WOP outcome criterion, for low EIRs, the TTI is 2-4 months for Burkina Faso and 2-6 months for Kenya, for medium EIRs, the TTI is 2-6 months for Burkina Faso and 4-8 months for Kenya, and for high EIRs, the TTI is 4-8 months for both Burkina Faso and Kenya. Low gene drive allele fitness costs and high effector gene efficacies result in TTIs approaching the low end of these ranges, while high gene drive allele fitness costs and low effector gene efficacies result in TTIs approaching the high end. Settings with higher EIRs are associated with higher TTIs because the mosquito population required to produce these EIRs is larger, and we consider unchanged release sizes, resulting in lower release ratios.

**Fig 4.**
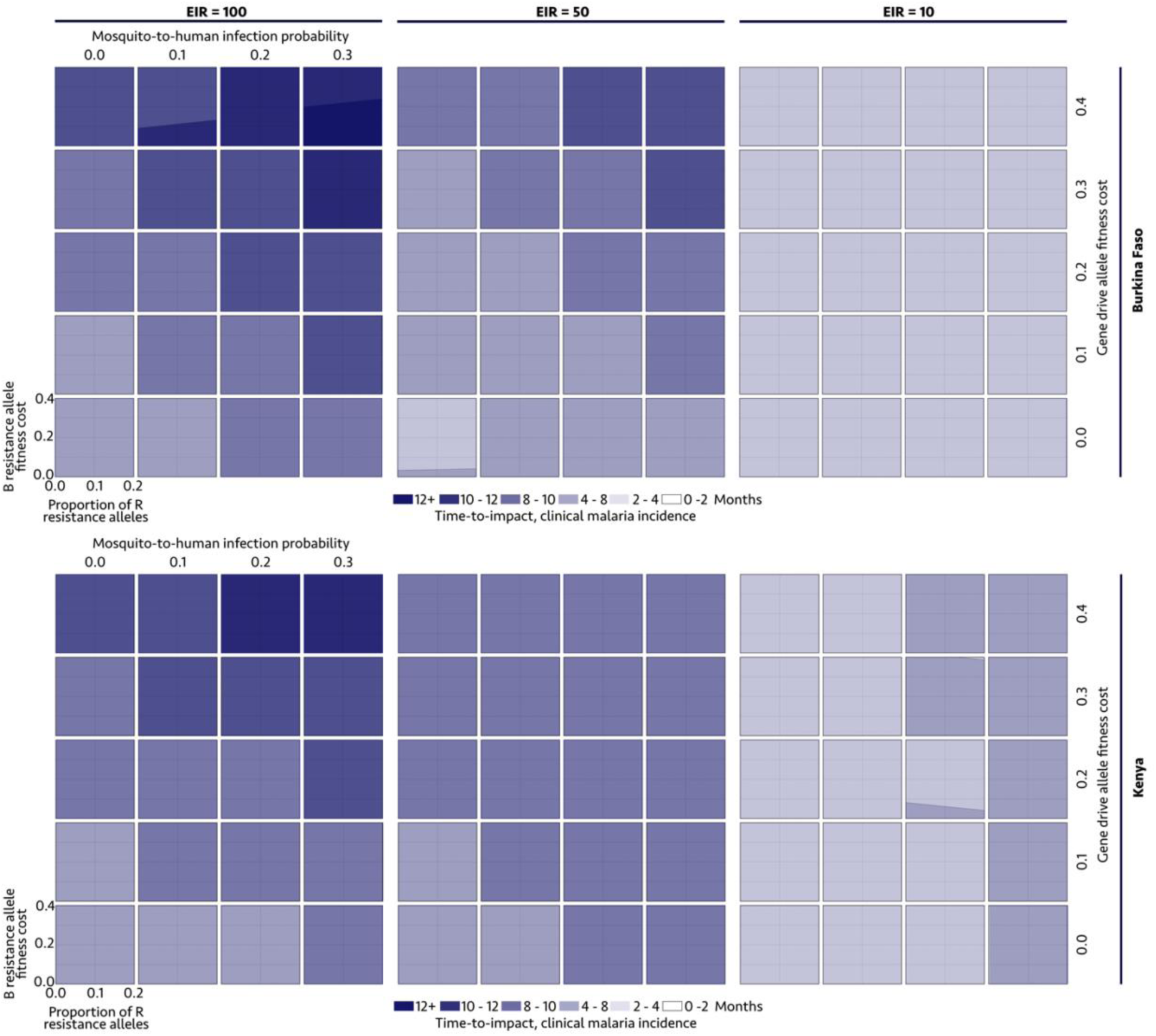
Gene drive parameter space satisfying defined times-to-impact (TTIs), i.e. times to a 50% reduction in clinical malaria incidence. TTIs are depicted for two country settings (Burkina Faso and Kenya, defined by their seasonal profile in **Fig 1B** and intervention coverage profile in **Table 1**) and three transmission settings (entomological inoculation rates, or EIRs, of 100, 50 and 10 infective bites per person per year). Gene drive parameters explored include: i) the fitness cost associated with being homozygous for the gene drive (H) allele, ii) the probability of mosquito-to-human transmission for mosquitoes having the H allele, iii) the fitness cost associated with being homozygous for the out-of-frame or otherwise costly B resistance allele, and iv) the proportion of generated resistance alleles that are in-frame and cost-free (R). The homing rate parameter is fixed at 0.95, since proposed outcome criteria were found to be insensitive to its value within a feasible range.

The TTI outcome criterion is satisfied for all regions of gene drive parameter space that satisfy WOP outcome criteria for the malaria-induced mortality metric (**Figs S5** of **S1 Text**), and for the malaria prevalence metric for low-to-medium EIRs (**Fig S6** of **S1 Text**). TTIs for the malaria-induced mortality metric closely reflect those for the clinical incidence metric, with modest reductions in TTI due to severe disease being suppressed slightly sooner than clinical disease following an intervention. For the malaria prevalence metric, TTIs exceed one year for high EIRs, reflecting the fact that reductions in prevalence take some time to manifest for low gene drive release ratios. TTIs for malaria prevalence are notably faster (2-8 months) for low-to-medium EIRs.

### Gene drive products are favored that generate fewer functional resistance alleles and non-functional resistance alleles that are more costly

The potential of a gene drive system to persist in a population for a sustained period of time, once most available wild-type alleles have been cleaved, is dependent on its ability to compete with drive-resistant alleles. In this sense, resistance allele parameters have a large impact on WOP criteria. To illustrate this, **Fig 5** depicts threshold values of *s_H_* and *b_H_* below which the clinical malaria incidence WOP exceeds three years, and above which it does not, for distinct sets of resistance allele parameters, *s_B_* and *p_R_*. Diagonal straight lines on this plot represent the trade-off between gene drive allele fitness cost and probability of mosquito-to-human infection in satisfying the WOP criterion. The movement of these lines upwards with decreasing values of the proportion of R alleles and/or increasing values of B allele fitness costs implies that, as cost-free (functional) resistance alleles are generated less frequently and as costly (non-functional) resistance alleles are associated with greater costs, greater drive allele fitness costs can be tolerated and/or lower effector gene efficacies. For instance, consider a Burkina Faso setting with an EIR of 100 infectious bites per person per year, a probability of transmission, *b_H_*, of 0.05, and a B allele fitness cost, *s_B_*, of 0.4. As the proportion of cost-free resistance alleles, *p_R_*, decreases from 0.2 to 0.1 to 0, the threshold H allele fitness cost, *s_H_*, satisfying the WOP criterion increases from 0.055 to 0.095 to 0.133. Alternatively, consider the same setting and probability of transmission, this time with the proportion of cost-free resistance alleles, *p_R_*, set to 0.1. Now as the B allele fitness cost, *s_B_*, increases from 0.1 to 0.2 to 0.4, the threshold H allele fitness cost, *s_H_*, satisfying the WOP criterion increases from 0.038 to 0.055 to 0.095.

**Fig 5.**
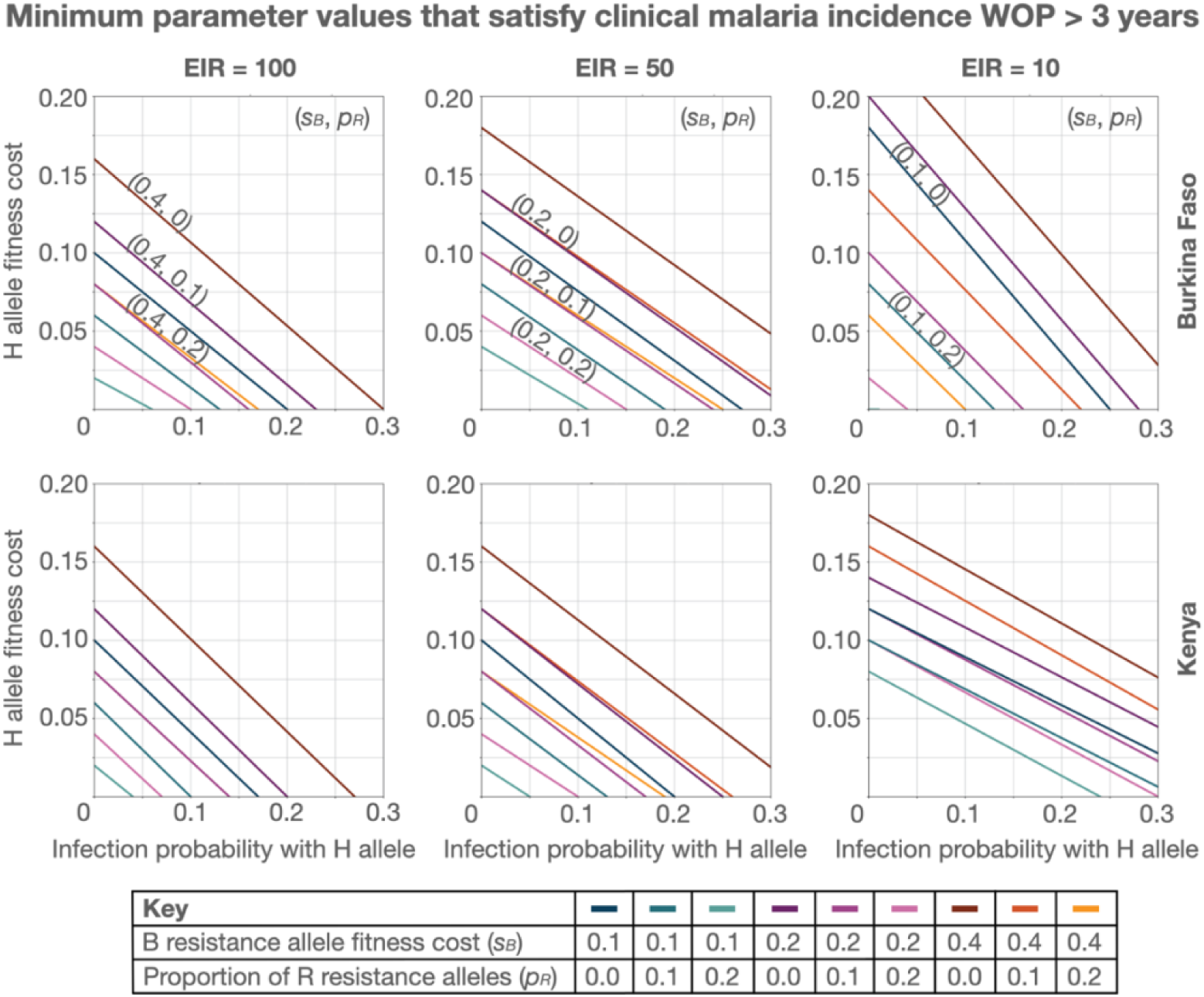
Minimum gene drive parameter values satisfying clinical malaria incidence window-of-protection greater than three years. Lines depict values of *s_H_* (H allele fitness cost) and *b_H_* (mosquito-to-human infection probability for mosquitoes having the H allele) below which the clinical malaria incidence window-of-protection exceeds three years, and above which it does not. Each line depicts a distinct set of resistance allele parameters -i.e., *s_B_* (B resistance allele fitness cost) and *p_R_* (proportion of R resistance alleles). The homing rate parameter, *h*, is fixed at 0.95. Threshold parameter values are depicted for two country settings (Burkina Faso and Kenya, defined by their seasonal profile in **Fig 1B** and intervention coverage profile in **Table 1**) and three transmission settings (entomological inoculation rates, or EIRs, of 100, 50 and 10 infective bites per person per year).

The same trends in higher tolerable *s_H_* and *b_H_* values for smaller *p_R_* values and higher *s_B_* values are seen for alternative outcome metrics. Threshold parameter values satisfying the malaria-induced mortality WOP criterion closely reflect thresholds for the clinical malaria incidence metric (**Fig S7** of **S1 Text**), likely due to the similarity in how mortality (a fraction of severe malaria incidence) and clinical malaria incidence are calculated. Threshold parameter values satisfying the malaria prevalence WOP criterion are more restrictive (**Fig S8** of **S1 Text**), especially for EIRs of 50-100 infective bites per person per year. This is again likely due to the delay in prevalence reductions being observed as a result of long-lasting untreated malaria infections.

### Target gene drive parameter values are interdependent

Results from **Figs 3-5** demonstrate that target gene drive parameter values are interdependent, i.e., the value of one parameter required to achieve a target outcome depends on the values of others. We therefore consider a range of assumptions and design options for potential gene drive constructs, and consider target parameters for each (**Table 2**). Given the relative insensitivity of target outcome criteria to the homing rate, *h*, we set this parameter to 0.95, reflecting constructs currently being considered for malaria vector control [4,5,32]. Second, given uncertainty surrounding fitness costs in the field, and especially regarding resistance alleles, we consider three scenarios for B allele fitness costs, *s_B_*: 0.1, 0.2 and 0.4. Third, for the proportion of resistance alleles that are in-frame/cost-free, *p_R_*, we consider a default value of ⅙. This is consistent with the estimated value for the Rec*kh* construct in *An. stephensi* [28], and with a reasonable estimate that about a third of mutations preserve the reading frame, and about half of those are cost-free. We also consider potential reductions in the value of *p_R_* achieved through guide RNA (gRNA) multiplexing, i.e., if multiple gRNAs within a drive construct target multiple nearby sequences within the target site, then the chance of generating in-frame/cost-free resistance alleles could be reduced multiplicatively [33,47]. We consider additional values of *p_R_* of (⅙)^2^ and (⅙)^3^ representing constructs having two and three gRNAs, respectively. Fourth, given the promise of current malaria-refractory effector genes available for *An. gambiae* [5,37], we consider mosquito-to-human infection probabilities, *b_H_*, of 0.01, 0.05 and 0.10. The probability of mosquito-to-human infection for a wild-type mosquito is estimated at 0.55 [38], so these represent infection-blocking efficacies of 98.1%, 90.9% and 81.8%, respectively. Finally, we consider an additional gene drive design in which the gene drive targets an essential gene required in at least one copy for mosquito viability [28,30]. This renders BB individuals unviable, while B allele heterozygotes are viable and have the same B allele fitness costs as for the default design. For each design and parameter set, we determine threshold values for H allele fitness cost, *s_H_*, that satisfy the target outcome of a clinical malaria incidence WOP exceeding three years.

**Table 2.**
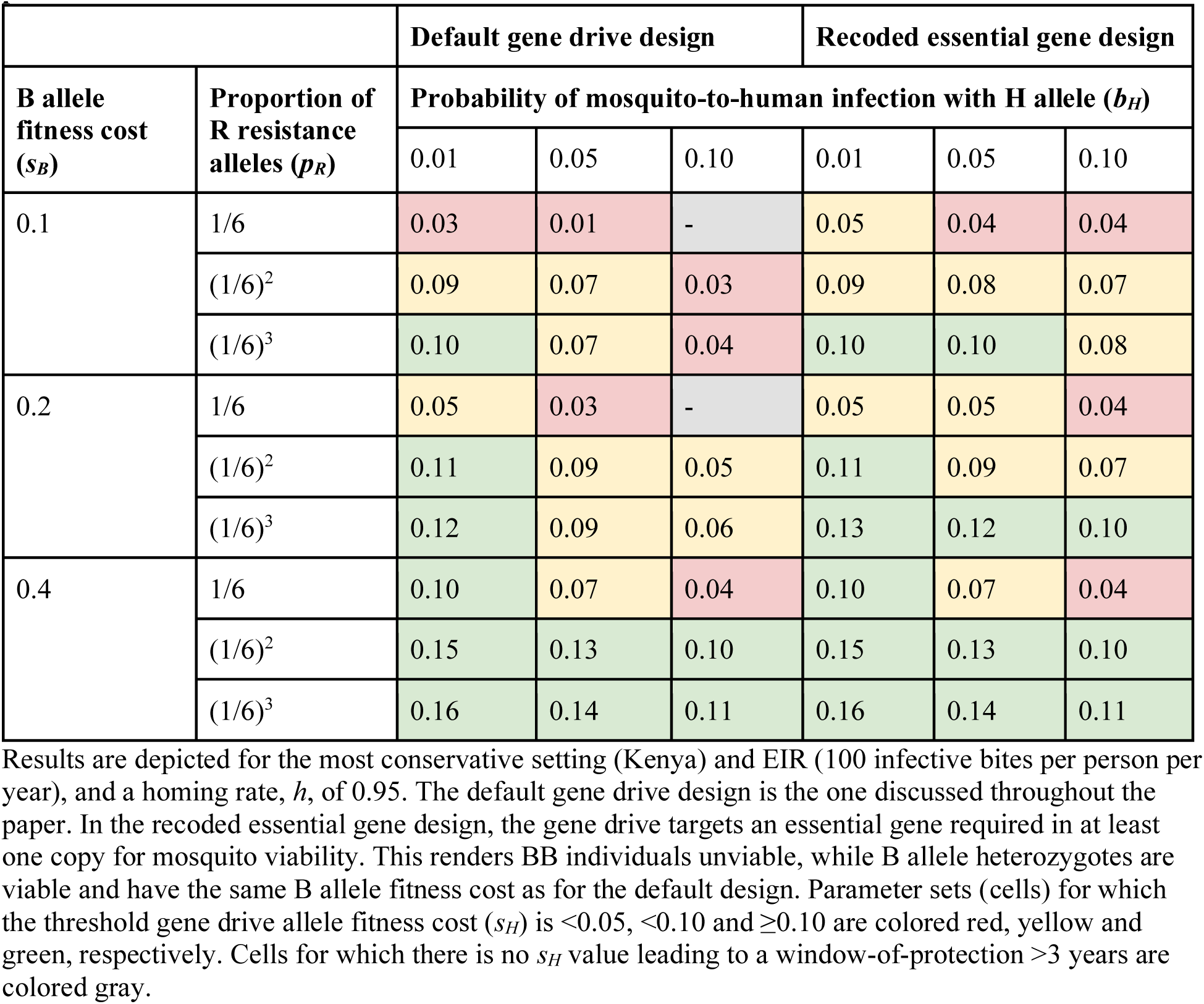
Threshold gene drive allele fitness cost (*s_H_*) satisfying clinical malaria incidence window-of-protection greater than three years.

|  |  | Default gene drive design |  |  | Recoded essential gene design |  |  |
| --- | --- | --- | --- | --- | --- | --- | --- |
| B allele fitness cost ($s_B$) | Proportion of R resistance alleles ($p_R$) | Probability of mosquito-to-human infection with H allele ($b_H$) | | | | | |
|  |  | 0.01 | 0.05 | 0.10 | 0.01 | 0.05 | 0.10 |
| 0.1 | 1/6 | 0.03 | 0.01 | - | 0.05 | 0.04 | 0.04 |
|  | (1/6)<sup>2</sup> | 0.09 | 0.07 | 0.03 | 0.09 | 0.08 | 0.07 |
|  | (1/6)<sup>3</sup> | 0.10 | 0.07 | 0.04 | 0.10 | 0.10 | 0.08 |
| 0.2 | 1/6 | 0.05 | 0.03 | - | 0.05 | 0.05 | 0.04 |
|  | (1/6)<sup>2</sup> | 0.11 | 0.09 | 0.05 | 0.11 | 0.09 | 0.07 |
|  | (1/6)<sup>3</sup> | 0.12 | 0.09 | 0.06 | 0.13 | 0.12 | 0.10 |
| 0.4 | 1/6 | 0.10 | 0.07 | 0.04 | 0.10 | 0.07 | 0.04 |
|  | (1/6)<sup>2</sup> | 0.15 | 0.13 | 0.10 | 0.15 | 0.13 | 0.10 |
|  | (1/6)<sup>3</sup> | 0.16 | 0.14 | 0.11 | 0.16 | 0.14 | 0.11 |

Results are depicted for the most conservative setting (Kenya) and EIR (100 infective bites per person per year), and a homing rate, $h$ , of 0.95. The default gene drive design is the one discussed throughout the paper. In the recoded essential gene design, the gene drive targets an essential gene required in at least one copy for mosquito viability. This renders BB individuals unviable, while B allele heterozygotes are viable and have the same B allele fitness cost as for the default design. Parameter sets (cells) for which the threshold gene drive allele fitness cost ( $s_H$ ) is $<0.05$ , $<0.10$ and $\geq 0.10$ are colored red, yellow and green, respectively. Cells for which there is no $s_H$ value leading to a window-of-protection $>3$ years are colored gray.

Results in **Table 2** reveal sets of parameter values for which the WOP criterion is satisfied for both the “default gene drive” and “recoded essential gene” designs. Due to interdependence of parameters, when the value of any specific parameter is enhanced (i.e., *s_H_*, *b_H_* or *p_R_* is decreased, or *s_B_* is increased), tolerable ranges for all other parameters are expanded. We consider minimally essential values of *b_H_*, *s_B_* and *p_R_* to be those for which the threshold value of *s_H_* is ≥0.10 (green cells in **Table 2**). This is motivated by the fact that an H allele fitness cost of 0.10 is already somewhat ambitious, particularly considering that reliable measurement is not possible prior to a field release, and hence it is best to err on the side of overestimating *s_H_* for a given construct. For both gene drive designs, ballpark values of *b_H_*, *s_B_* and *p_R_* that satisfy this threshold condition are: i) a probability of mosquito-to-human infection with the H allele, *b_H_*, of ≤0.05, ii) a B allele fitness cost, *s_B_*, of ≥0.20, and iii) a proportion of R resistance alleles, *p_R_*, of ≤(⅙)^2^. While the recoded essential gene design does not improve upon the minimally essential parameter values, it does expand tolerable ranges for these parameters, given permissive values of other parameters. E.g., in the case where *s_B_*=0.20 and *p_R_*=(⅙)^3^, a *b_H_* value of 0.10 is tolerable for the recoded essential gene design, while a *b_H_* value of 0.05 is on the cusp of being tolerable for the default gene drive design.

Finally, results in **Table S2** of **S1 Text** align with conclusions throughout the manuscript regarding alternative outcome metrics. For the malaria-induced mortality metric, minimally essential parameter values reflect those for the clinical malaria incidence metric; while for the malaria prevalence metric, minimally essential parameter values are much more restrictive: i) a probability of mosquito-to-human infection with the H allele, *b_H_*, of ≤0.01, ii) a B allele fitness cost, *s_B_*, of ≥0.40, and iii) a proportion of R resistance alleles, *p_R_*, of ≤(⅙)^2^.

## 4. Discussion

Model-informed TPPs for gene drive mosquito products are especially important given that the epidemiological target outcomes of interest can only be observed following a release [13], and such a release may be difficult to remediate [10]. We modeled and simulated a broad range of gene drive product parameters for releases in two country settings and at three malaria transmission levels. This allowed us to characterize regions of gene drive parameter space expected to satisfy target outcome criteria: a 50% reduction in clinical malaria incidence, a rate of spread that would produce this impact in less than a year, and a duration of impact of at least three years. We found that the TTI criterion was always satisfied when the WOP criterion was satisfied, allowing us to focus on the latter. We also explored alternative outcome metrics, reductions in malaria-induced mortality and prevalence. Reductions in malaria-induced mortality mirrored those for clinical incidence, while the prevalence metric led to overly restrictive outcome criteria, likely due to delays in reductions in prevalence resulting from enduring untreated infections.

Regarding gene drive product parameters, we explored rates of homing and resistance allele generation, fitness costs associated with the gene drive and out-of-frame/costly resistance alleles, and the efficacy of the effector gene at reducing mosquito-to-human transmission. Simulations support the conclusion that, for feasible parameter ranges, the WOP is notably least influenced by the homing rate, and is most influenced by fitness costs associated with the gene drive allele, and the efficacy of the effector gene. A trade-off between homing allele fitness cost and effector gene efficacy is apparent in which, for high effector gene efficacies, larger homing allele fitness costs can be accommodated. Resistance allele parameters are also highly influential on target outcomes, as they determine how long the gene drive allele persists in the population after most available wild-type alleles have been cleaved. If fewer functional resistance alleles are generated, and if non-functional resistance alleles have larger fitness costs, then a wider range of effector gene efficacies and homing allele fitness costs can be accommodated. We also find that a gene drive design that renders BB individuals unviable expands the range of allowable parameter values in some cases. Since target parameter values are interdependent, it is only possible to specify approximate TPP thresholds (ideal and minimally essential) for each. We discuss these thresholds for each parameter separately in the following paragraphs.

### Fitness of gene drive and resistance alleles are highly influential on outcome criteria, but difficult to measure prior to a release

In the interests of parsimony, we considered just two fitness parameters in our analysis, *s_H_* and *s_B_*, which represent fitness costs associated with being homozygous for the H and B alleles, respectively. Fitness costs were assumed to be additive. Of note, *s_H_* represents fitness costs associated with both the gene drive and linked effector gene. Outcomes of interest were most sensitive to *s_H_*, while *s_B_* was the third most important parameter. This presents a conundrum for assessing product readiness according to a TPP since, while the performance of a gene drive product is highly dependent on these parameters, fitness is difficult, if not impossible, to reliably measure prior to a field release. Nevertheless, this provides an incentive for gene drive developers to focus engineering efforts on minimizing gene drive fitness costs, while maximizing costs on non-functional resistance alleles. An approach that seeks to address both criteria is to have the gRNA target site be an essential gene, while including a copy of this gene within the drive construct. This approach was employed for the Rec*kh* construct in *An. stephensi*, for which BB females were rendered unviable, while laboratory measurements indicated no fitness cost on the H allele [28]. The approach has also been employed in *Drosophila*, described as the home-and-rescue (HomeR) design, with a similar laboratory fitness profile [48]. Modeling results suggest additional fitness costs attributed to B allele heterozygotes will have added benefits for H allele/effector gene persistence. Other approaches to minimizing H allele fitness cost include: i) using promoters that restrict Cas9 expression to the germline, hence having little effect on somatic tissue [28,32], ii) using blood meal-inducible promoters that restrict antimalarial gene expression [5], and iii) choosing gRNA target sites having minimal fitness consequences, although this latter approach may also reduce fitness costs associated with B alleles [5].

Based on our analysis, we recommend H allele fitness costs in homozygotes, *s_H_*, ≤0.10 (minimally essential) and ≤0.05 (ideal), and B allele fitness costs in homozygotes, *s_B_*, ≥0.20 (minimally essential) and ≥0.40 (ideal). Given our assumptions, this implies fitness costs of ≤0.05 (minimally essential) and ≤0.025 (ideal) for H allele heterozygotes, and ≥0.10 (minimally essential) and ≥0.20 (ideal) for B allele heterozygotes. Higher H allele costs and lower B allele costs are tolerable given permissive values of *h*, *b_H_* and *p_R_* in our model. Importantly, these targets should be interpreted as fitness costs once the introduced alleles have introgressed into the wild genetic background and shed fitness effects associated with the genetic background of the release strain. This post-introgression fitness should reflect the competitive dynamics between drive and drive-resistant alleles once the majority of wild-type alleles in the population have been cleaved, while higher release strain fitness costs prior to introgression may be offset through supplemental releases.

### Efficacy of the effector gene(s) is highly influential, and some insight can be gained from lab measurements

Efficacy of the effector gene(s) was the second most important parameter in predicting TTI and WOP for all three outcome metrics. This parameter is difficult to measure directly, adding to the conundrum of assessing product readiness prior to a field release. For malaria vaccine and acquired immunity studies, human challenge experiments with infected mosquitoes are routinely performed [49,50]; however, to our knowledge, this has not yet been done as part of the development pathway for population modification gene drive mosquitoes. That said; we can gain some insight into effector gene efficacy from laboratory studies. E.g., for the AgTP13 (*An. gambiae*) and AcTP13 (*An. coluzzii*) population modification strains, dual effector genes are included that encode monoclonal antibodies targeting parasite ookinetes and sporozoites [5]. Reductions in sporozoite numbers in the salivary glands of infected mosquitoes heterozygous or homozygous for this construct were measured using a long-cultured laboratory *P. falciparum* strain, and the data support potential threshold-dependent transmission-blocking [5]. In another approach to engineering malaria-refractory effectors, a midgut gene of *An. gambiae* was augmented to secrete two antimicrobial peptides previously shown to interfere with *Plasmodium* development [37]. In this case, infection experiments were performed using both *P. falciparum* and a rodent malaria model system parasite, *P. berghei*, and sporozoites were shown to be both delayed in development and reduced in number alongside reductions in oocyst diameter [37].

Delays and reductions in sporozoite counts cannot be directly translated into reductions in mosquito-to-human malaria infection; however, significant delays and/or reductions are suggestive of highly effective transmission-blocking. Infection experiments in mice with *Plasmodium yolei* reveal a nonlinear relationship between sporozoite load and infection probability, with a threshold model and threshold of ∼10,000 sporozoites providing the best fit [51]. While the value of this threshold will vary between species (i.e., parasite, vector and host), this finding suggests that low (i.e., sub-threshold) sporozoite counts may indeed result in very low mosquito-to-human transmission probabilities. Given the importance of this parameter in satisfying TTI and WOP outcome criteria, we encourage continued refinement of malaria-refractory effector genes to further lower sporozoite loads and infection probabilities. We recommend a mosquito-to-human infection probability with the H allele, *b_H_*, of ≤0.05 (minimally essential) or ≤0.01 (ideal), which represents a >90% (minimally essential) or >98% (ideal) reduction in transmission probability compared to wild-type. That said; higher infection probabilities can be tolerated given permissive values of *h*, *s_H_*, *s_B_* and *p_R_*, as was demonstrated in previous modeling of AgTP13 and AcTP13 [5].

### Rates of homing and resistance allele generation are measurable, but less influential

Ironically, of the five product parameters that we explored, the two that are most well studied and measurable, the rates of homing and resistance allele generation, are also the two that least influence the outcome criteria. That said; measures of influence depend on the range of parameter values explored, so for the homing rate, this is partly a reflection of the high rates of accurate HDR achieved for recently engineered population modification gene drives in *Anopheles* [4,5,27,28,32]. These values are invariably >0.95, so we conservatively explored values between 0.8 and 1; but even with this expanded range, the inheritance bias associated with the H allele is sufficient to drive it into a population within the timeframe of a field trial. We therefore conclude that several candidate population modification gene drive systems in *Anopheles* [5,32] have already satisfied TPP requirements for the homing rate parameter, and development efforts should now shift to optimizing resistance allele, fitness and effector gene efficacy parameters. We recommend a homing rate, *h*, ≥0.95 (minimally essential and ideal); however lower values, possibly even below 0.8, are tolerable given permissive values of *s_H_*, *s_B_*, *p_R_* and *b_H_*. We recommend this value due to it already having been achieved, and because we used a value of 0.95 in generating our targets for other parameters, which depend to some extent on the value of *h*.

The proportion of resistance alleles that are in-frame/cost-free (R) was the fourth most important parameter (of five) in predicting the outcome criteria. Naturally, as fewer R alleles are generated, it takes longer for them to outcompete H alleles after most available wild-type alleles have been cleaved, leading the drive system and linked malaria-refractory genes to persist in the population for longer. Several approaches have been proposed to reduce the rate of resistance allele generation, most commonly, gRNA multiplexing [33,47], and in some cases, male-specific Cas expression [33]. We recommend a proportion of R resistance alleles, *p_R_*, ≤(⅙)^2^ (minimally essential) and ≤(⅙)^3^ (ideal). This represents a default *p_R_* value of ⅙, consistent with about a third of mutations preserving the reading frame, and about half of those being cost-free, and two (minimally essential) or three (ideal) gRNAs acting independently in disrupting the target site. This is a parameter that can be measured through laboratory crosses and sequencing, and hence the number of gRNAs required should be tailored according to empirical measurements rather than theory. As for all other product parameters, larger values of *p_R_* are tolerable given permissive values of *h*, *s_H_*, *s_B_* and *b_H_*. A draft, model-informed TPP summarizing considerations for all five explored gene drive product parameters is depicted in **Table 3**.

**Table 3.** Model-inferred target product parameter values for population modification gene drive systems.

| Product parameter | Minimally essential value | Ideal value | Notes |
| --- | --- | --- | --- |
| Probability of accurate homology-directed repair given cleavage ($h$) | $\geq 0.95$ | $\geq 0.95$ | Adequate value already achieved |
| Proportion of resistant alleles that are in-frame/cost-free ($p_R$) | $\leq (\frac{1}{6})^2$ | $\leq (\frac{1}{6})^3$ | ~2 guide RNAs (minimally essential), ~3 guide RNAs (ideal) |
| Probability of mosquito-to-human infection with H allele ($b_H$) | $\leq 0.05$ | $\leq 0.01$ | >90% reduction (minimally essential), >98% reduction (ideal) |
| H allele fitness cost in homozygotes ($s_H$) | $\leq 0.10$ | $\leq 0.05$ | Heterozygote fitness costs are half these values |
| B allele fitness cost in homozygotes ($s_B$) | $\geq 0.20$ | $\geq 0.40$ | Heterozygote fitness costs are half these values |

### Comparison to other modeling studies

Results from this analysis are broadly consistent with other studies [20,52], albeit with differences due to modeling frameworks and assumptions. The most similar study to ours, that of Leung *et al.* [20], explores characteristics of population modification gene drives that lead to malaria elimination in a Sahelian setting for a variety of EIRs (10, 30 and 80 infectious bites per person per year). Similar to our study, theirs argues that homing rates for current population modification gene drives in *Anopheles* are adequately high (>0.95), and that attention in gene drive development should now focus on effector gene efficacy and fitness costs. They argue for a transmission-blocking efficacy ≥90% (which equates to an infection probability in our model ≤0.055), as this leads to malaria elimination for realistic parameter values given a moderate EIR.

Our results and those of Leung *et al.* [20] diverge regarding fitness costs. In the model of Leung *et al.* [20], gene drive allele fitness costs upwards of 0.40 can be tolerated provided effector gene efficacy is sufficiently high. This is partly due to the manner in which fitness costs are modeled. We consider reductions in female fecundity and male mating competitiveness, while they consider increases in adult mortality rate. High fitness costs in their model therefore lead to reduced mosquito population density and hence could contribute to reductions in malaria transmission. The other difference is structural; the model of Leung *et al.* [20] is stochastic, with human and mosquito populations being scaled down by a factor of 100 to enable tractable individual-based simulation. These features facilitate malaria elimination as a modeled outcome. In contrast, the malaria transmission model in our framework [22] is deterministic, represented by a set of differential equations, and hence malaria elimination is not captured. Our key outcome of interest, the WOP, is more dependent on the relative fitness of gene drive and resistance alleles, which determine the duration that malaria-refractory genes persist in the population. Given these assumptions, our model advocates for a much smaller gene drive allele fitness cost of ≤0.10. This comparison highlights the sensitivity of model predictions to model structure and assumptions, particularly for highly influential parameters such as fitness costs.

A separate analysis by Beaghton *et al.* [52] captures the competition between gene drive and resistance alleles over a time period similar to ours. Their study doesn’t explicitly include a malaria transmission model, instead modeling reduction in vectorial capacity as a malaria-refractory effector gene spreads into a population, and exploring how the duration of reduction in vectorial capacity varies with a range of gene drive parameters. Direct comparison is difficult, as molecular processes and fitness profiles are modeled differently. Beaghton *et al.* [52] model loss of the effector and/or nuclease separately, through either the homing process or spontaneous mutation, and separate fitness costs are associated with the nuclease and effector genes; however, durations of protection for default parameter values are broadly consistent. In the analysis by Leung *et al.* [20], parameters describing resistance allele generation do not vary, while a parameter describing pre-existing drive-resistance in the population is varied instead.

### Modeling framework and limitations

A general concern with gene drive modeling is that parsimonious models may overestimate the potential field impact of a tool that has not yet been field-tested due to: i) simplifications in the description of the ecosystem, and ii) limitations in quantifying rare molecular processes that may have a large impact on the outcome of an intervention at-scale. Perhaps the most significant ecosystem simplification in gene drive models to date [17,20,22] is that only one mosquito species is modeled, implicitly assuming there are no other species present that contribute to malaria transmission in a significant way. This may be the case in some locations; but is not the norm in mainland Africa. In the Sahel, *An. gambiae*, *An. coluzzi* and *Anopheles arabiensis* all vector malaria and vary in relative seasonal abundance throughout the year [53], while in east Africa, *An. arabiensis* and *Anopheles funestus* are often present at varying degrees depending on season and geography [54]. Modeling a single species when other species are available to vector the pathogen will overestimate intervention impact; however, reliable models of multi-species dynamics remain to be developed and will inevitably be tied to the location of the data to which they are fitted. This is an important area of future research. Another important ecosystem-level limitation to our analysis is the lack of a spatial component. Our model may be seen as representing a panmictic population akin to a field trial site. Spatial analysis by Beaghton *et al.* [52] demonstrates how, as resistance alleles accumulate, the effector gene reaches a lower maximum frequency and has a shorter duration of protection at larger distances from the release site. A solution to this would be to model seeding of the drive system throughout a landscape, as Leung *et al.* [20] did.

In the interests of parsimony, our model ignores molecular outcomes expected to occur at slow rates as a result of homing or mutation processes. These include loss of the effector gene through the homing process, or loss of the nuclease, effector gene or target site through spontaneous mutation [52]. Another possible outcome is evolution of effector-resistance within the parasite, effectively decreasing the efficacy of the effector as resistance spreads [55]. These processes are likely less influential over the short timescale modeled here (six years following a release). Regarding fitness costs associated with gene drive alleles, as mentioned earlier, we have modeled these as being associated with reductions in female fecundity and male mating competitiveness. This is supported by laboratory fitness assays for candidate drive systems [5,32], although lifespan reduction has also been observed [5], and comparative modeling demonstrates this to be important in determining model outcomes [20]. Given the importance of fitness cost parameters, further field study is encouraged, whether this be through highly confined releases, or releases of self-limiting or effector-only constructs.

## 5. Conclusion

Here, we used a mathematical model to provide a model-informed TPP for population modification gene drive systems. Simulated product parameters included rates of homing and R resistance allele generation, fitness costs of the drive allele and B resistance alleles, and efficacy of the malaria-refractory gene(s). Simulated seasonal settings included Burkina Faso and Kenya, each at three transmission intensities (high, medium and low). Target outcome criteria included a >50% reduction in clinical malaria incidence for a duration of >3 years (WOP criterion), and a time-to-impact of <1 year (TTI criterion). For reasonable parameter values, we found that, if the WOP criterion is satisfied, the TTI criterion is automatically satisfied too. The most important parameters for satisfying the WOP criterion are (in order): i) fitness cost of the drive allele, ii) effector gene efficacy, iii) fitness cost of the B resistance allele, iv) proportion of generated resistance alleles that are R, and v) homing rate. A trade-off exists between the fitness cost of the drive allele and effector gene efficacy, i.e., for higher effector gene efficacies, larger drive allele fitness costs can be tolerated. In the intermediate term, once most wild-type alleles have been cleaved, persistence of the drive allele is largely dependent on selective competition between the drive and resistance alleles. Low rates of R resistance allele generation are therefore preferred, and achievable through the use of multiple gRNAs. More costly B resistance alleles will allow the drive allele to outcompete them. Homing rates already achieved in *Anopheles* do not need to be improved upon. A conundrum exists that the most important product parameters for predicting field efficacy are those that can only be reliably measured in the field. Nevertheless, these parameters are most important for ongoing development efforts: low drive allele fitness cost, high B resistance allele fitness cost, and high effector gene efficacy. We recommend prioritizing their refinement, and making measurements via laboratory proxies and/or confined field study, where possible.

## Supporting information

**Table S1.** Parameter values describing mosquito bionomics, vector control and malaria epidemiology.

**S1 Text.** Results for alternative outcome metrics. Feature importance and product parameters satisfying target outcome criteria for malaria-induced mortality and malaria prevalence metrics.

## Data availability statement

Simulation code is available on GitHub at: https://github.com/amondal2/ReplacementTPP.

## Acknowledgements

We thank Dr. Anthony James and Dr. George Dimopoulos for comments on the manuscript.

