## Supplementary material for "A model-informed target product profile for population modification gene drives for malaria control": S1 Text


**Results for alternative outcome metrics**

Here, we depict target product profile analysis results - including feature importance, and regions of gene drive parameter space that satisfy target outcome criteria - for alternative outcome metrics. The two alternative outcome metrics are all-ages malaria prevalence, and malaria-induced mortality. For each simulation, these are generated as a time-series, and two target outcomes are derived: i) window-of-protection, which measures the duration for which the outcome metric is below 50% its seasonal mean, and ii) time-to-impact, which measures the time from initial release to the outcome metric falling to 50% its seasonal mean. Malaria prevalence, also referred to as the *Plasmodium falciparum* parasite rate (*PfPR*), refers to the proportion of the human population that harbors the malaria pathogen, regardless of symptoms or treatment status. For the Imperial College London (ICL) malaria model [1,2], this is given by,

$$PfPR(t) = \sum_{a \in A} (A_a(t) + U_a(t) + T_a(t) + D_a(t))/N_H \ . \qquad \text{EqS1}$$

Here, $N_H$ represents the total human population size, and $A_a(t)$, $U_a(t)$, $T_a(t)$ and $D_a(t)$ represent the number of people in age group $a$ who are either asymptomatic but detectable by rapid diagnostic test (RDT), asymptomatic and undetectable by RDT, symptomatically infected and treated, or symptomatically infected and untreated (diseased), respectively, at time $t$. Secondly, for the ICL model [1,2], malaria-induced mortality is proportional to the incidence of severe malaria at time $t$, and is given by,

$$Deaths(t) = \nu \sum_{a \in A} \lambda_{H,a}(t)\, \theta_a(t)\, (S_a(t) + A_a(t) + U_a(t)) \ . \qquad \text{EqS2}$$

Here, $\lambda_{H,a}(t)$ represents the force of infection on humans (probability of infection per person per unit time) for age group $a$ at time $t$, $\theta_a(t)$ represents the probability of acquiring severe disease upon infection for age group $a$ at time $t$, $S_a(t)$ represents the number of people in age group $a$ who are susceptible at time $t$, and $\nu$ represents the probability of death for a case of severe disease. The derivation and parameterization of this formula is provided in Griffin *et al.* [2]. Results for these alternative outcome metrics follow.

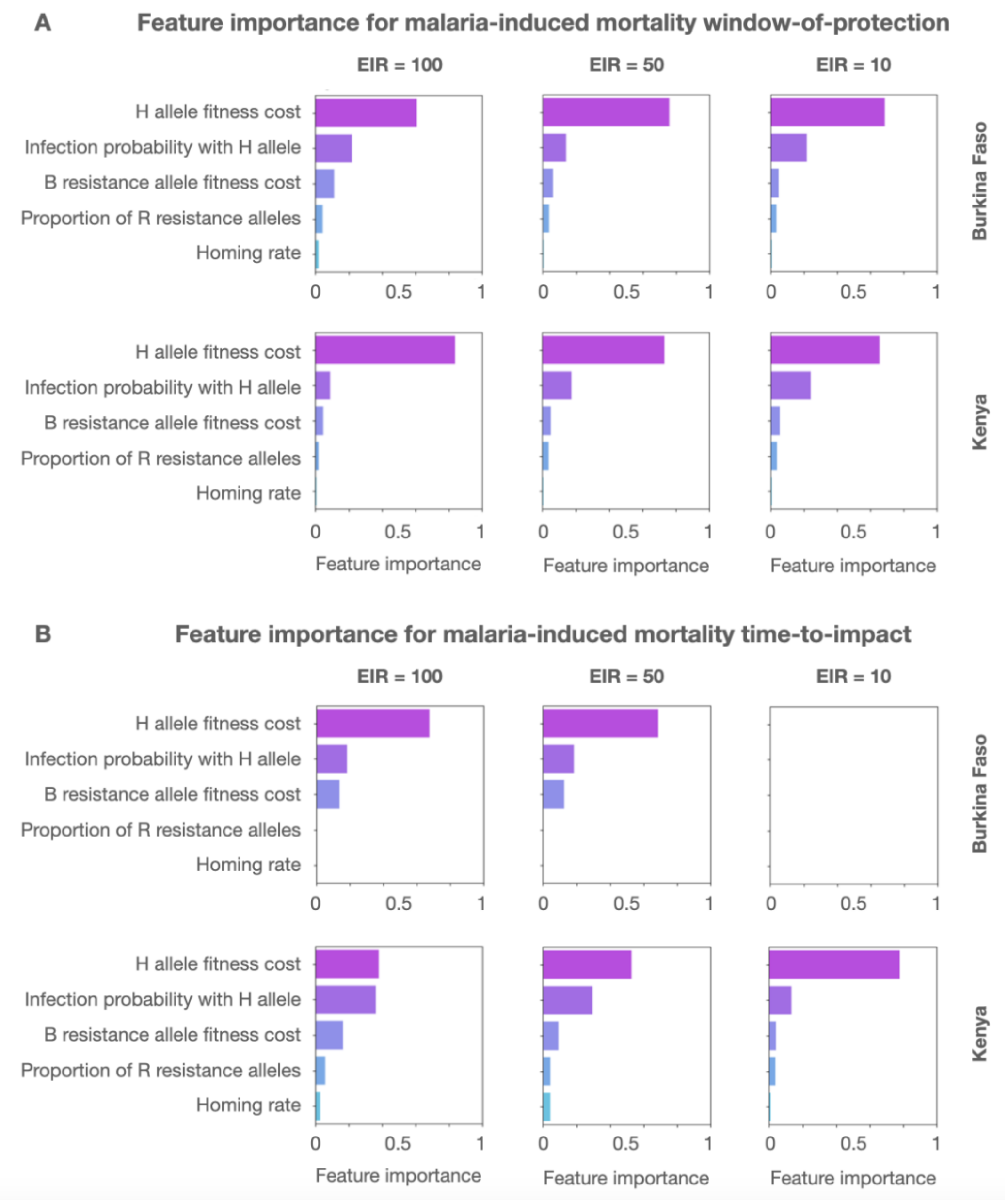


**Fig S1. Feature importance of gene drive product parameters (malaria-induced mortality outcome).** Permutation feature importance values are depicted for gene drive product parameters for two country settings (Burkina Faso and Kenya), three transmission intensities (entomological inoculation rates of 100, 50 and 10 per person per year), and two target outcomes - **(A)** window-of-protection (i.e., the duration for which malaria prevalence is below 50% its seasonal mean), and **(B)** time-to-impact (i.e., the time from initial release to malaria prevalence falling to 50% its seasonal mean). Parameters explored include: i) the fitness cost associated with being homozygous for the gene drive (H) allele, ii) the probability of mosquito-to-human transmission for mosquitoes having the H allele with linked antimalarial effector gene(s), iii) the fitness cost associated with being homozygous for the out-of-frame or otherwise costly B resistance allele, iv) the proportion of generated resistance alleles that are in-frame and cost-free (R), and v) the homing rate, or rate of accurate homology-directed repair given cleavage. Permutation feature importance is calculated on a 0-1 scale.

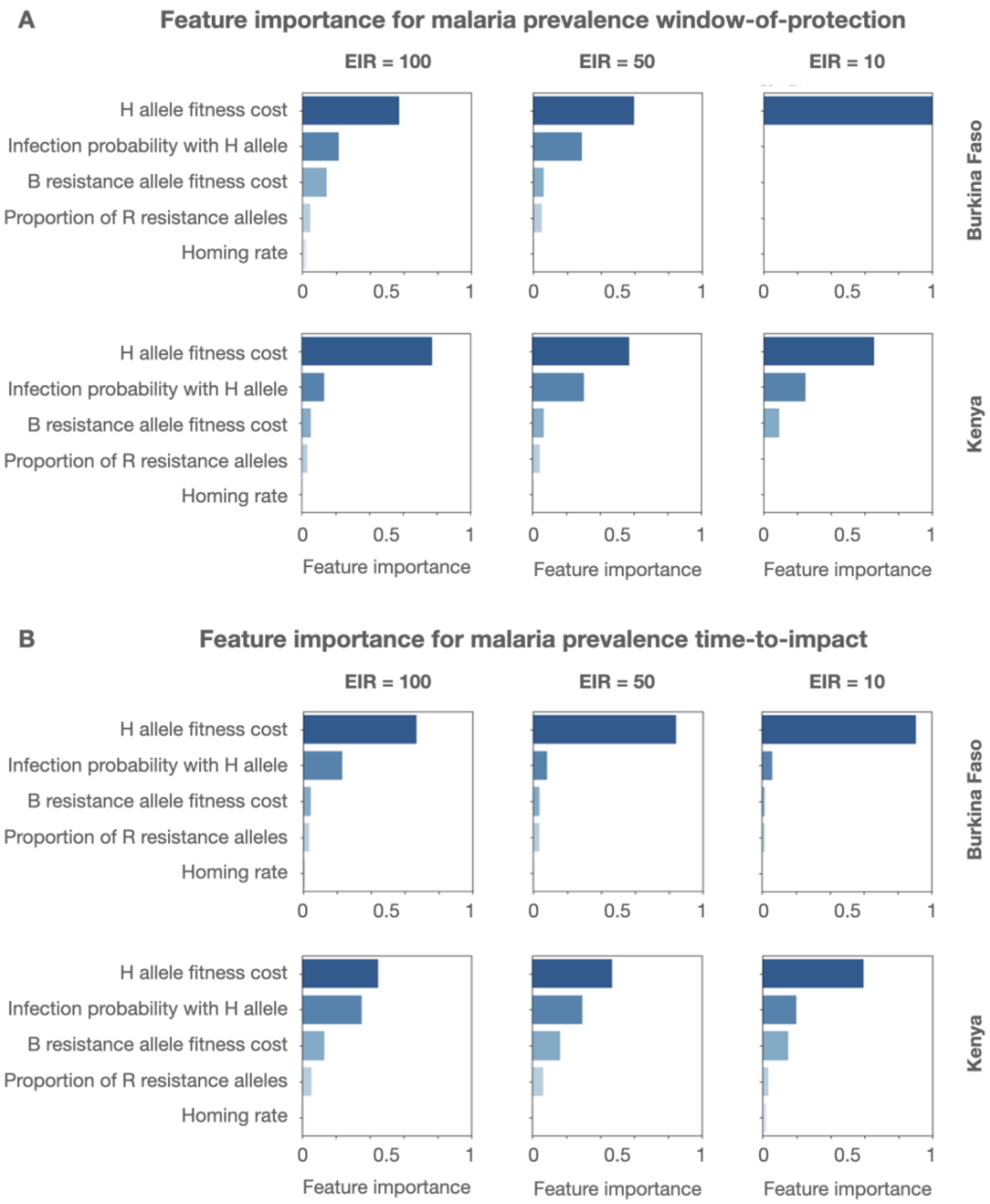


**Fig S2. Feature importance of gene drive product parameters (malaria prevalence outcome).** Permutation feature importance values are depicted for gene drive product parameters for two country settings (Burkina Faso and Kenya), three transmission intensities (entomological inoculation rates of 100, 50 and 10 per person per year), and two target outcomes - **(A)** window-of-protection (i.e., the duration for which malaria-induced mortality is below 50% its seasonal mean), and **(B)** time-to-impact (i.e., the time from initial release to malaria-induced mortality falling to 50% its seasonal mean). Parameters explored include: i) the fitness cost associated with being homozygous for the gene drive (H) allele, ii) the probability of mosquito-to-human transmission for mosquitoes having the H allele with linked antimalarial effector gene(s), iii) the fitness cost associated with being homozygous for the out-of-frame or otherwise costly B resistance allele, iv) the proportion of generated resistance alleles that are in-frame and cost-free (R), and v) the homing rate, or rate of accurate homology-directed repair given cleavage. Permutation feature importance is calculated on a 0-1 scale.

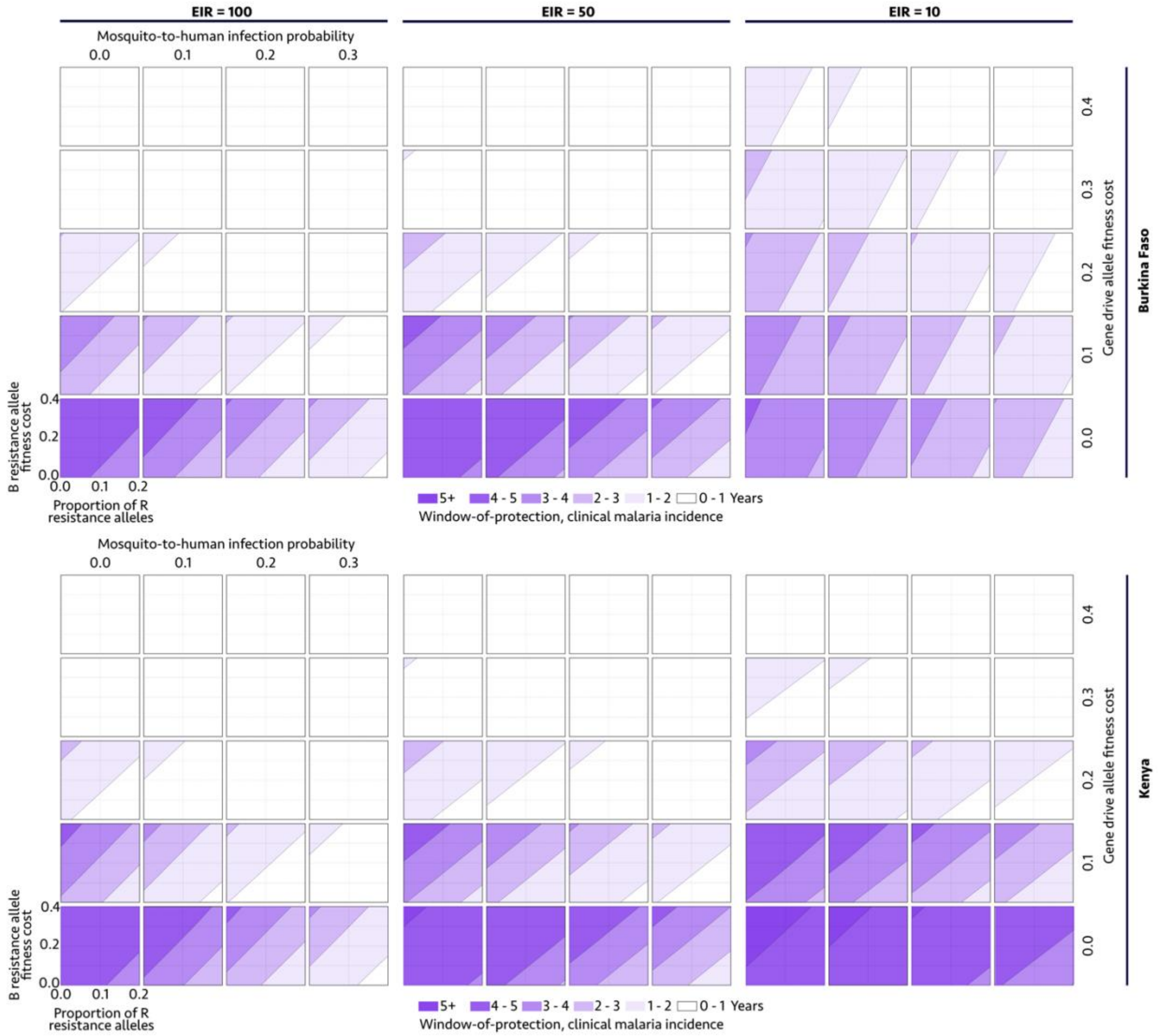


**Fig S3. Gene drive parameter space satisfying a >50% reduction in malaria-induced mortality for defined durations (i.e., window-of-protection, or WOP).** WOPs are depicted for two country settings (Burkina Faso and Kenya, defined by their seasonal profile in **Fig 1B** and intervention coverage profile in **Table 1**) and three transmission settings (entomological inoculation rates, or EIRs, of 100, 50 and 10 infective bites per person per year). Gene drive parameters explored include: i) the fitness cost associated with being homozygous for the gene drive (H) allele, ii) the probability of mosquito-to-human transmission for mosquitoes having the H allele, iii) the fitness cost associated with being homozygous for the out-of-frame or otherwise costly B resistance allele, and iv) the proportion of generated resistance alleles that are in-frame and cost-free (R). The homing rate parameter is fixed at 0.95, since proposed outcome criteria were found to be insensitive to its value within a feasible range.

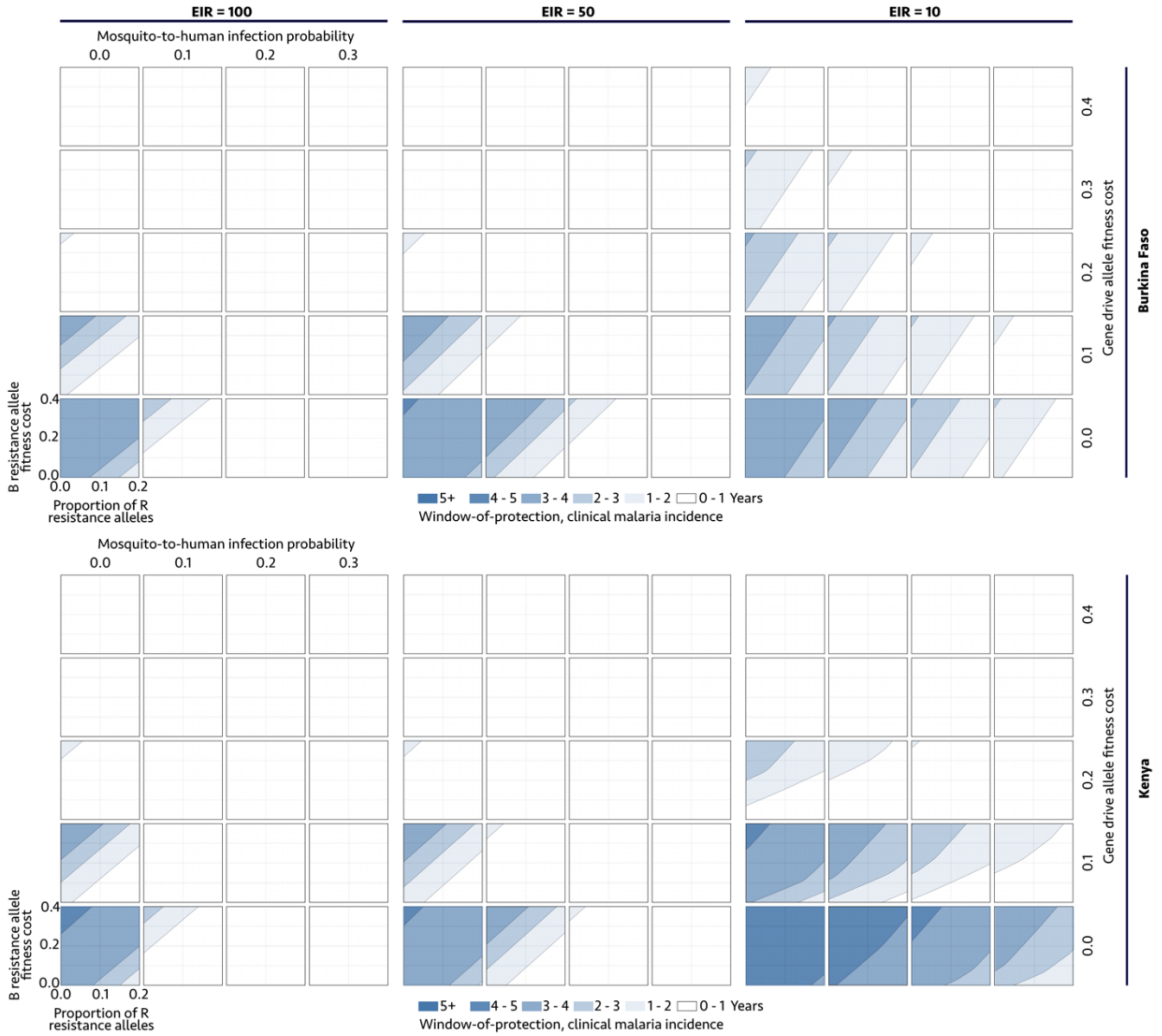


**Fig S4. Gene drive parameter space satisfying a >50% reduction in malaria prevalence for defined durations (i.e., window-of-protection, or WOP).** WOPs are depicted for two country settings (Burkina Faso and Kenya, defined by their seasonal profile in **Fig 1B** and intervention coverage profile in **Table 1**) and three transmission settings (entomological inoculation rates, or EIRs, of 100, 50 and 10 infective bites per person per year). Gene drive parameters explored include: i) the fitness cost associated with being homozygous for the gene drive (H) allele, ii) the probability of mosquito-to-human transmission for mosquitoes having the H allele, iii) the fitness cost associated with being homozygous for the out-of-frame or otherwise costly B resistance allele, and iv) the proportion of generated resistance alleles that are in-frame and cost-free (R). The homing rate parameter is fixed at 0.95, since proposed outcome criteria were found to be insensitive to its value within a feasible range.

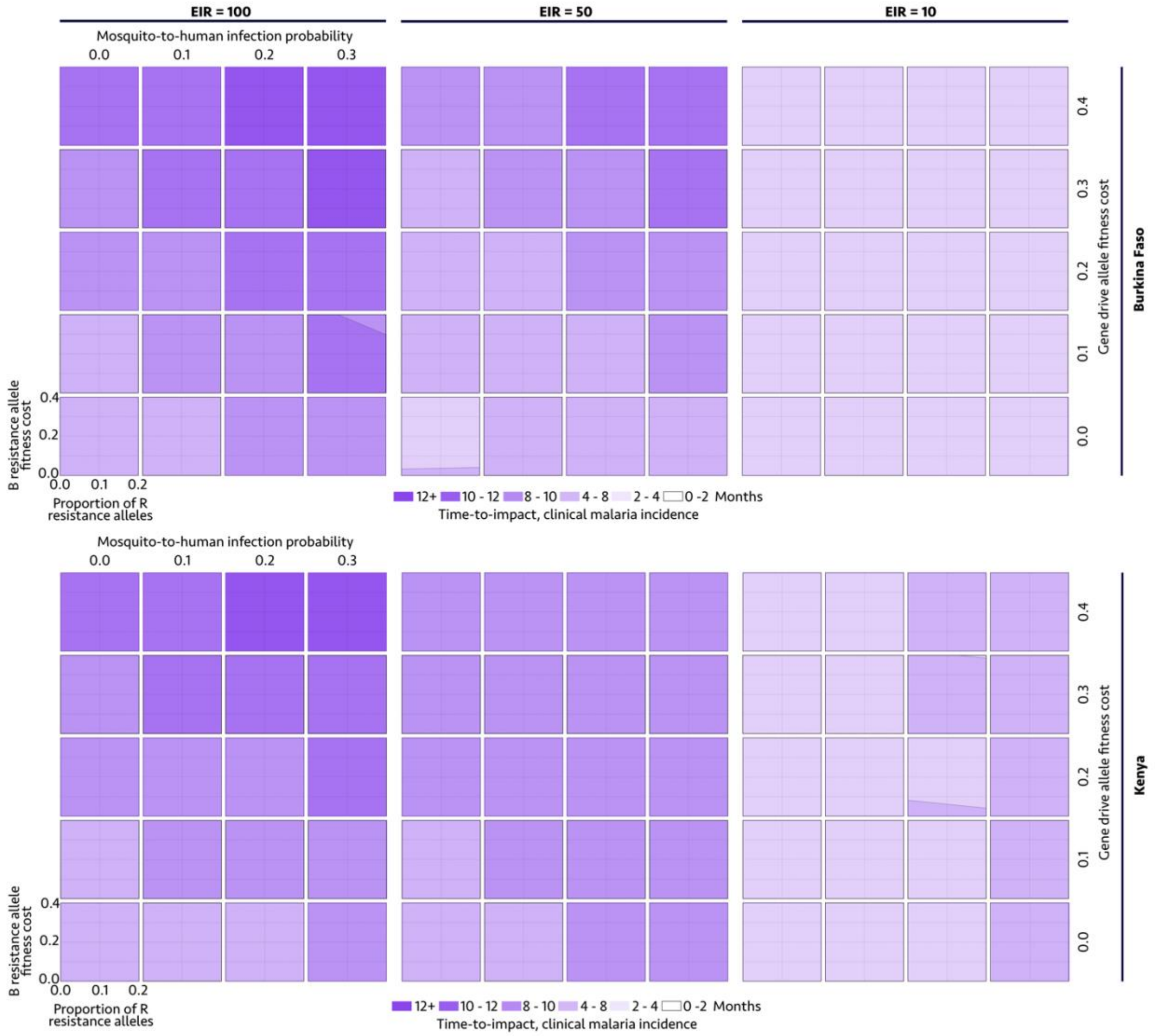


**Fig S5. Gene drive parameter space satisfying defined times to impact (TTIs), i.e. times to a 50% reduction in malaria-induced mortality.** TTIs are depicted for two country settings (Burkina Faso and Kenya, defined by their seasonal profile in **Fig 1B** and intervention coverage profile in **Table 1**) and three transmission settings (entomological inoculation rates, or EIRs, of 100, 50 and 10 infective bites per person per year). Gene drive parameters explored include: i) the fitness cost associated with being homozygous for the gene drive (H) allele, ii) the probability of mosquito-to-human transmission for mosquitoes having the H allele, iii) the fitness cost associated with being homozygous for the out-of-frame or otherwise costly B resistance allele, and iv) the proportion of generated resistance alleles that are in-frame and cost-free (R). The homing rate parameter is fixed at 0.95, since proposed outcome criteria were found to be insensitive to its value within a feasible range.

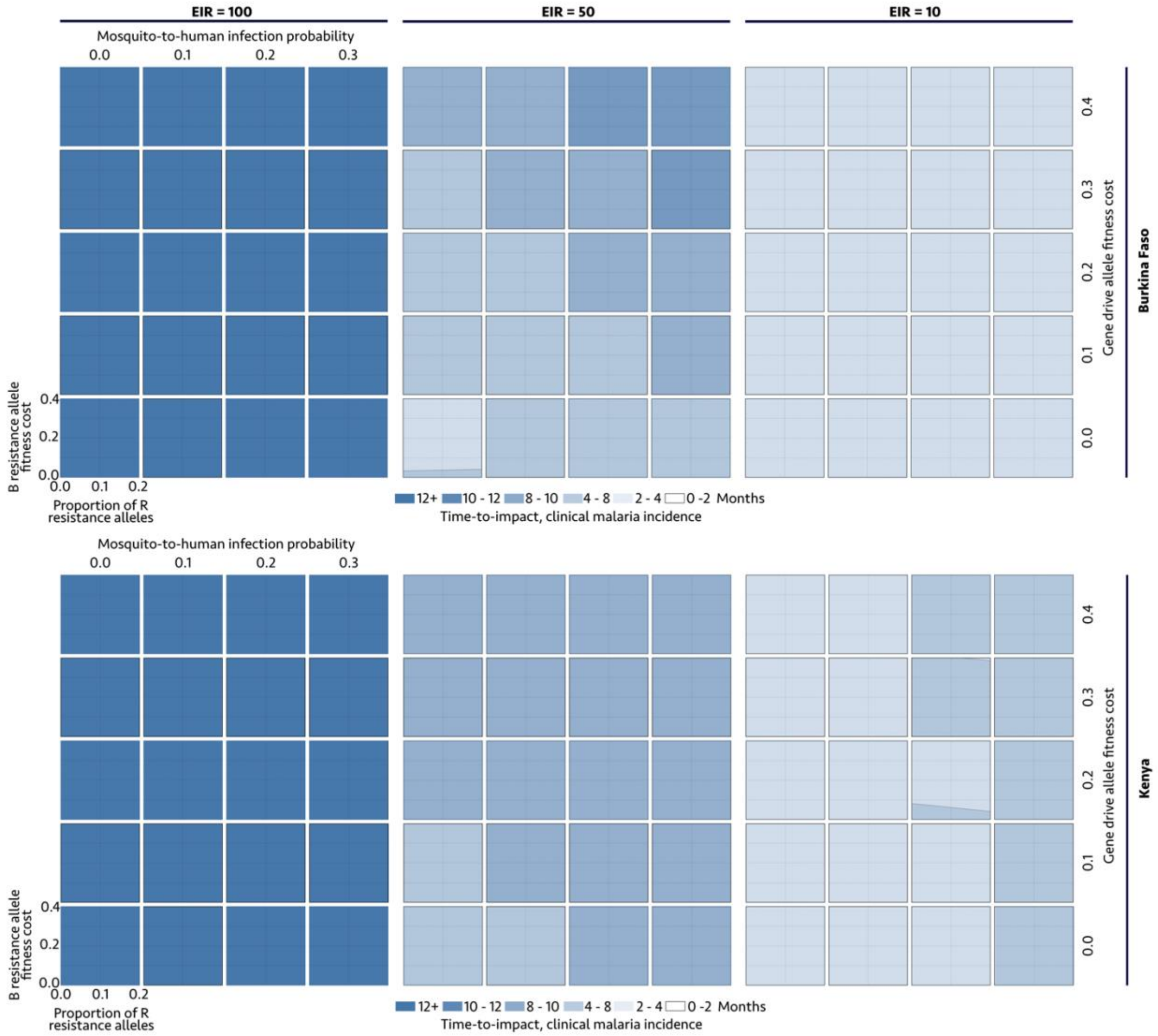


**Fig S6. Gene drive parameter space satisfying defined times to impact (TTIs), i.e. times to a 50% reduction in malaria prevalence.** TTIs are depicted for two country settings (Burkina Faso and Kenya, defined by their seasonal profile in **Fig 1B** and intervention coverage profile in **Table 1**) and three transmission settings (entomological inoculation rates, or EIRs, of 100, 50 and 10 infective bites per person per year). Gene drive parameters explored include: i) the fitness cost associated with being homozygous for the gene drive (H) allele, ii) the probability of mosquito-to-human transmission for mosquitoes having the H allele, iii) the fitness cost associated with being homozygous for the out-of-frame or otherwise costly B resistance allele, and iv) the proportion of generated resistance alleles that are in-frame and cost-free (R). The homing rate parameter is fixed at 0.95, since proposed outcome criteria were found to be insensitive to its value within a feasible range.

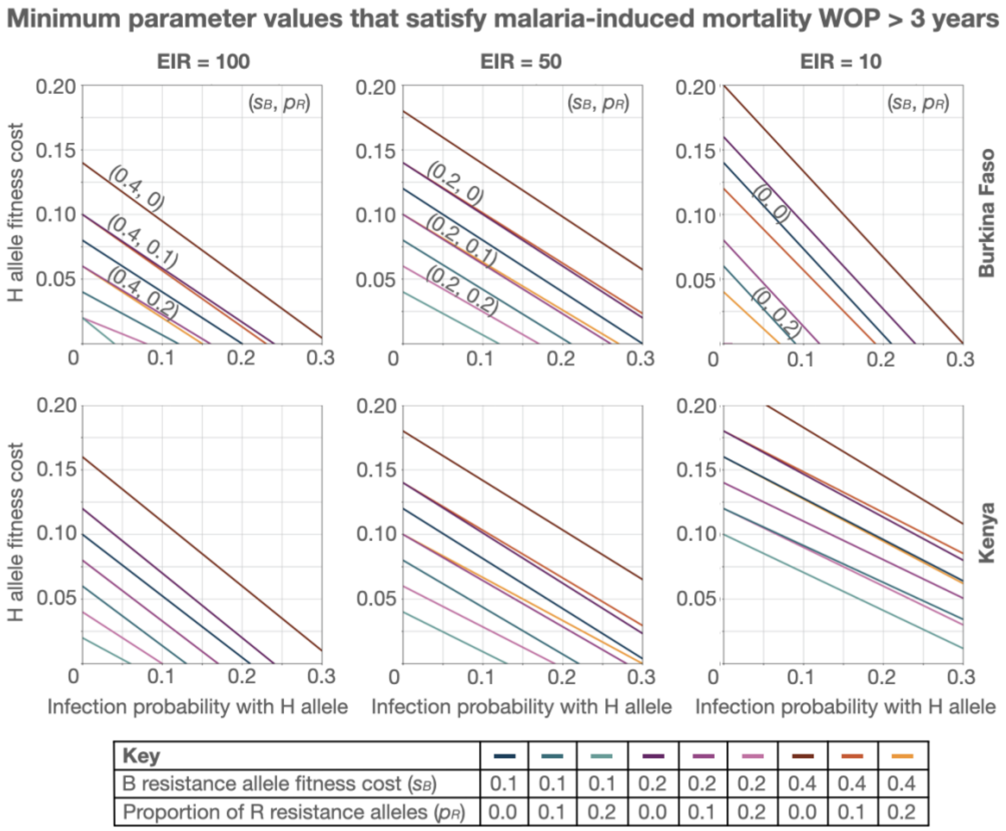


**Fig S7. Minimum gene drive parameter values satisfying malaria-induced mortality window-of-protection greater than three years.** Lines depict values of $s_H$ (H allele fitness cost) and $b_H$ (mosquito-to-human infection probability for mosquitoes having the H allele) below which the malaria-induced mortality window-of-protection exceeds three years, and above which it does not. Each line depicts a distinct set of resistance allele parameters - i.e., $s_B$ (B resistance allele fitness cost) and $p_R$ (proportion of R resistance alleles). The homing rate parameter, $h$, is fixed at 0.95. Threshold parameter values are depicted for two country settings (Burkina Faso and Kenya, defined by their seasonal profile in **Fig 1B** and intervention coverage profile in **Table 1**) and three transmission settings (entomological inoculation rates, or EIRs, of 100, 50 and 10 infective bites per person per year).

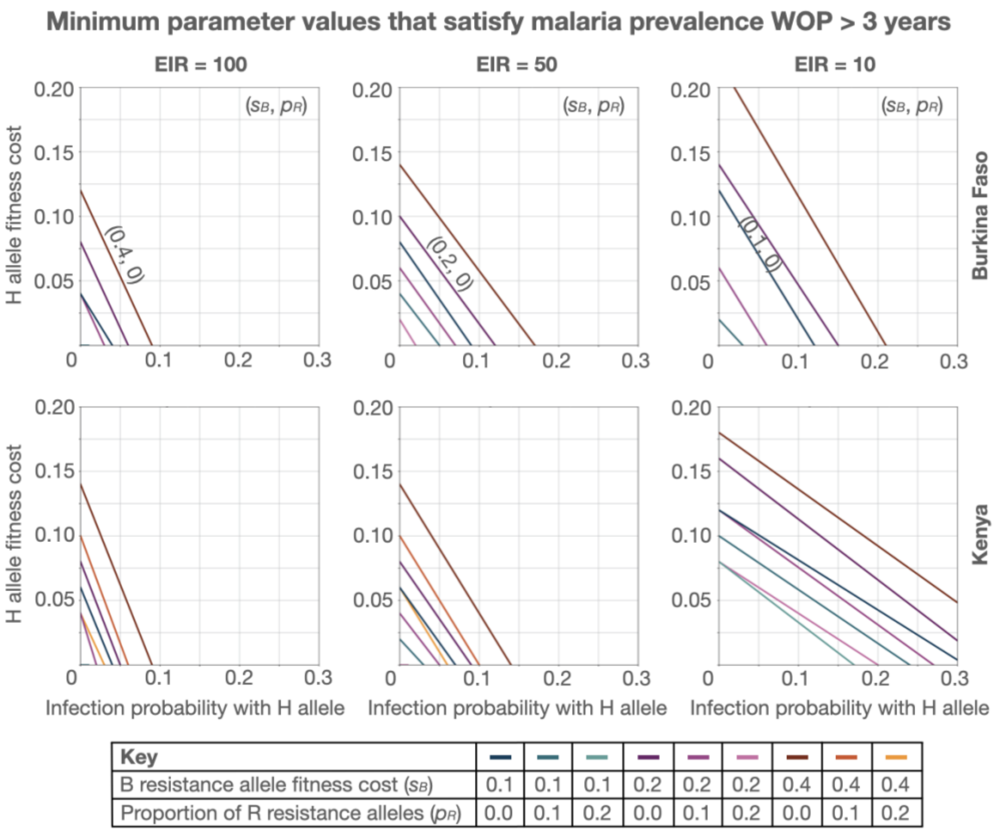


**Fig S8. Minimum gene drive parameter values satisfying malaria prevalence window-of-protection greater than three years.** Lines depict values of $s_H$ (H allele fitness cost) and $b_H$ (mosquito-to-human infection probability for mosquitoes having the H allele) below which the malaria prevalence window-of-protection exceeds three years, and above which it does not. Each line depicts a distinct set of resistance allele parameters - i.e., $s_B$ (B resistance allele fitness cost) and $p_R$ (proportion of R resistance alleles). The homing rate parameter, $h$, is fixed at 0.95. Threshold parameter values are depicted for two country settings (Burkina Faso and Kenya, defined by their seasonal profile in **Fig 1B** and intervention coverage profile in **Table 1**) and three transmission settings (entomological inoculation rates, or EIRs, of 100, 50 and 10 infective bites per person per year).

**Table S2. Threshold gene drive allele fitness cost ($s_H$) satisfying malaria-induced mortality and malaria prevalence windows of protection greater than three years.**

| | | **Malaria-induced mortality outcome metric** | | | **Malaria prevalence outcome metric** | | |
|---|---|---|---|---|---|---|---|
| **B allele fitness cost ($s_B$)** | **Proportion of R resistance alleles ($p_R$)** | **Probability of mosquito-to-human infection with H allele ($b_H$)** | | | | | |
| | | 0.01 | 0.05 | 0.10 | 0.01 | 0.05 | 0.10 |
| 0.1 | 1/6 | 0.04 | 0.02 | - | - | - | - |
| | $(1/6)^2$ | 0.10 | 0.08 | 0.05 | 0.04 | - | - |
| | $(1/6)^3$ | 0.11 | 0.08 | 0.06 | 0.05 | 0.00 | - |
| 0.2 | 1/6 | 0.06 | 0.04 | 0.01 | 0.00 | - | - |
| | $(1/6)^2$ | 0.13 | 0.10 | 0.08 | 0.07 | 0.00 | - |
| | $(1/6)^3$ | 0.13 | 0.10 | 0.08 | 0.08 | 0.01 | - |
| 0.4 | 1/6 | 0.10 | 0.08 | 0.05 | 0.06 | 0.00 | - |
| | $(1/6)^2$ | 0.15 | 0.13 | 0.11 | 0.12 | 0.06 | - |
| | $(1/6)^3$ | 0.16 | 0.14 | 0.12 | 0.13 | 0.07 | 0.01 |

Results are depicted for the most conservative setting (Kenya) and EIR (100 infective bites per person per year), and a homing rate, *h*, of 0.95. The default gene drive design is the one discussed throughout the paper. Parameter sets (cells) for which the threshold gene drive allele fitness cost ($s_H$) is $<0.05$, $<0.10$ and $\geq 0.10$ are colored red, yellow and green, respectively. Cells for which there is no $s_H$ value leading to a window-of-protection >3 years are colored gray.
