## Supplementary material for "A model-informed target product profile for population modification gene drives for malaria control": Table S1

**Table S1. Parameter values describing mosquito bionomics, vector control and malaria epidemiology.**

| Symbol | Parameter | Value | Reference |
|---|---|---|---|
| **Mosquito bionomics** | | | |
| $\beta$ | Egg production per adult female (per day) | 21 | [1] |
| $T_E$ | Mean duration of egg stage (days) | 3 | [1] |
| $T_L$ | Mean duration of larval stage (days) | 7 | [1] |
| $T_P$ | Mean duration of pupal stage (days) | 1 | [1] |
| $\mathrm{CV}(T_E)$ | Coefficient of variation, egg stage | 0.2 | [2] |
| $\mathrm{CV}(T_L)$ | Coefficient of variation, larval stage | 0.3 | [2] |
| $\mathrm{CV}(T_P)$ | Coefficient of variation, pupal stage | 0.2 | [2] |
| $K$ | Larval carrying capacity | Time-varying | [3] |
| $\mu$ | Adult mosquito mortality rate | Time-varying | [3] |
| $f$ | Blood feeding rate | 1/3 | [4] |
| $Q$ | Human blood index | 0.9 | [4] |
| **Vector control** | | | |
| $\theta_B$ | Bites taken on humans while they are in bed as a proportion of all bites taken on humans | 0.89 | [5,6] |
| $\theta_I$ | Bites taken on humans while they are indoors as a proportion of all bites taken on humans | 0.97 | [5,6] |
| $r_{LLIN}$ | Probability of repeating a feeding attempt in the presence of long-lasting insecticide-treated nets | 0.56 | [5,6] |
| $r_{IRS}$ | Probability of repeating a feeding attempt in the presence of indoor residual spraying | 0.60 | [5,6] |
| $s_{LLIN}$ | Probability of feeding and surviving in the presence of long-lasting insecticide-treated nets | 0.03 | [5,6] |
| $s_{IRS}$ | Probability of feeding and surviving in the presence of indoor residual spraying | 0 | [5,6] |
| **Malaria epidemiology** | | | |
| $N_H$ | Human population size | 1,000 | |
| $v$ | Proportion of severe malaria cases that result in death | 0.215 | [7] |
